# Alterations in chromatin accessibility and conformation elucidate genetic mechanisms in ASD

**DOI:** 10.1101/2025.04.02.25325129

**Authors:** Jiani Yin, Jerry Huang, Jing Ou, George Chen, Lucy K. Bicks, Brie Wamsley, Yuyan Cheng, Lawrence Chen, Jillian R. Haney, Rain Wong, Daniel H. Geschwind

## Abstract

Genetic risk for psychiatric disorders lies largely within non-coding regions, where the lack of detailed knowledge of gene regulation and chromatin structure has hampered understanding of disease mechanisms. We analyzed chromatin accessibility and 3D genome architecture in brains from 53 ASD and neurotypical individuals, including patients with (dup) 15q11-13. We observed reduced CTCF binding, which had dual effects: a) decreased chromatin accessibility at distal enhancers and downregulation of synaptic and neuronal target genes, and b) weakened TAD boundaries linked to DNA hypermethylation, impacting a distinct set of genes. These changes were associated with brain mQTLs, caQTLs, and rare variants increasing ASD risk, a subset of which we validated by CRISPR editing, supporting a causal relationship. Our analyses suggest that genetic variants contribute to risk in part through a combination of epigenetic changes, including disruption of distal enhancer accessibility and 3D genome organization in both idiopathic and a syndromic form of ASD.

## Introduction

Autism spectrum disorder (ASD) is a prevalent neurodevelopmental disorder (CDC^1^) characterized by deficits in social communication and interaction, and restricted or repetitive behaviors or interests, with symptom onset during early childhood^2^. ASD is highly heritable^3,4^. Its genetic architecture is complex and multifaceted, with rare, primarily *de novo* single gene mutations and copy number variants (CNVs) accounting for approximately 15-20% of cases^5^, and the cumulative effect of common genetic variants predicted to account for at least 50%^4,6^. Approximately 30-40% of the attributable genetic risk is unaccounted for^4,6^, but some is likely imparted by rare inherited variation^7–9^. Over 200 high-confidence ASD risk genes have been identified and they have been shown to converge on multiple biological processes, including synaptic, transcriptional, and chromatin remodeling pathways^8,10–13^.

Despite the substantial phenotypic and genetic heterogeneity observed in ASD, transcriptomic profiling of post-mortem brain reveals consistent molecular changes in the brain in a majority of autistic individuals, which have been validated and refined over time^11,14–18^. These alterations in mRNA and miRNA involve down-regulation of neuronal pathways that center on synaptic signaling and mitochondrial function, and up-regulation of multifaceted neural-immune responses in glia. Furthermore, attenuation of typical regional differences in gene expression between brain regions has been repeatedly observed in ASD^15,16,19^. Subsequent DNA methylation and histone acetylation studies have demonstrated that the up-regulated immune processes are associated with DNA hypomethylation, whereas promoter regions of down-regulated neuronal genes are frequently hyperacetylated, which may be compensatory^20,21^. Nevertheless, how these multiple epigenetic layers interact and how they might reflect causal genetic variation contributions to ASD remain largely unknown.

Advances in chromatin biology and genomics have yielded fundamental insights into gene regulatory mechanisms over the last decade^22–24^. The ability to measure chromatin accessibility, through techniques such as ATAC-seq (Assay for Transposase-Accessible Chromatin using sequencing), facilitates the identification of regulatory regions and associated transcription factors, as well as the characterization of chromatin structure dynamics^25^. The 3D genome, which is typically profiled by Hi-C, establishes connections between distal regulatory regions and their target genes^26,27^. Combining these methods, we profiled cerebral cortex from 53 ASD cases and controls, including 7 cases with a syndromic form of ASD, (dup) 15q11-13 (referred to as Dup15q syndrome hereafter). Our extensive dataset enabled us to investigate changes in chromatin accessibility and 3D chromosome conformation in ASD, linking these changes to transcriptomic dysregulation. We further identified transcription factor networks and genetic variants associated with these changes, providing causal links potentially underlying the pathophysiology of ASD. These findings provide a framework for deciphering genetic mechanisms by elucidating how genetic risk factors alter the transcriptional regulatory landscape and contribute to ASD etiology.

## Results

### Characterization of chromatin accessibility changes in ASD

We performed two genome-wide assays to interrogate chromatin structure in post-mortem brain from 19 individuals with idiopathic ASD and 19 age- and sex-matched neurotypical subjects, ATAC-seq which identifies open accessible regions, and Hi-C which links regulatory regions to their cognate target genes (Fig. 1a). In addition, we generated technical replicates for 8 of these samples, yielding a total of 46 ATAC-seq libraries, to ensure data reproducibility and control for technical variability. After extensive quality control (Methods, Supplementary Figure 1), the final analysis dataset comprised 38 ATAC-seq libraries from 13 ASD and 19 neurotypical subjects (Methods, Supplementary Table 1 and Supplementary Figure 1). We identified 128,036 reproducible open chromatin regions across the pooled dataset with an average peak length of 1,184 bp. In line with the previous studies^28,29^, over 15% of the ATAC peaks were located at gene promoters, 44% within gene bodies and 35% were intergenic (Fig. 1b).

**Fig. 1.**
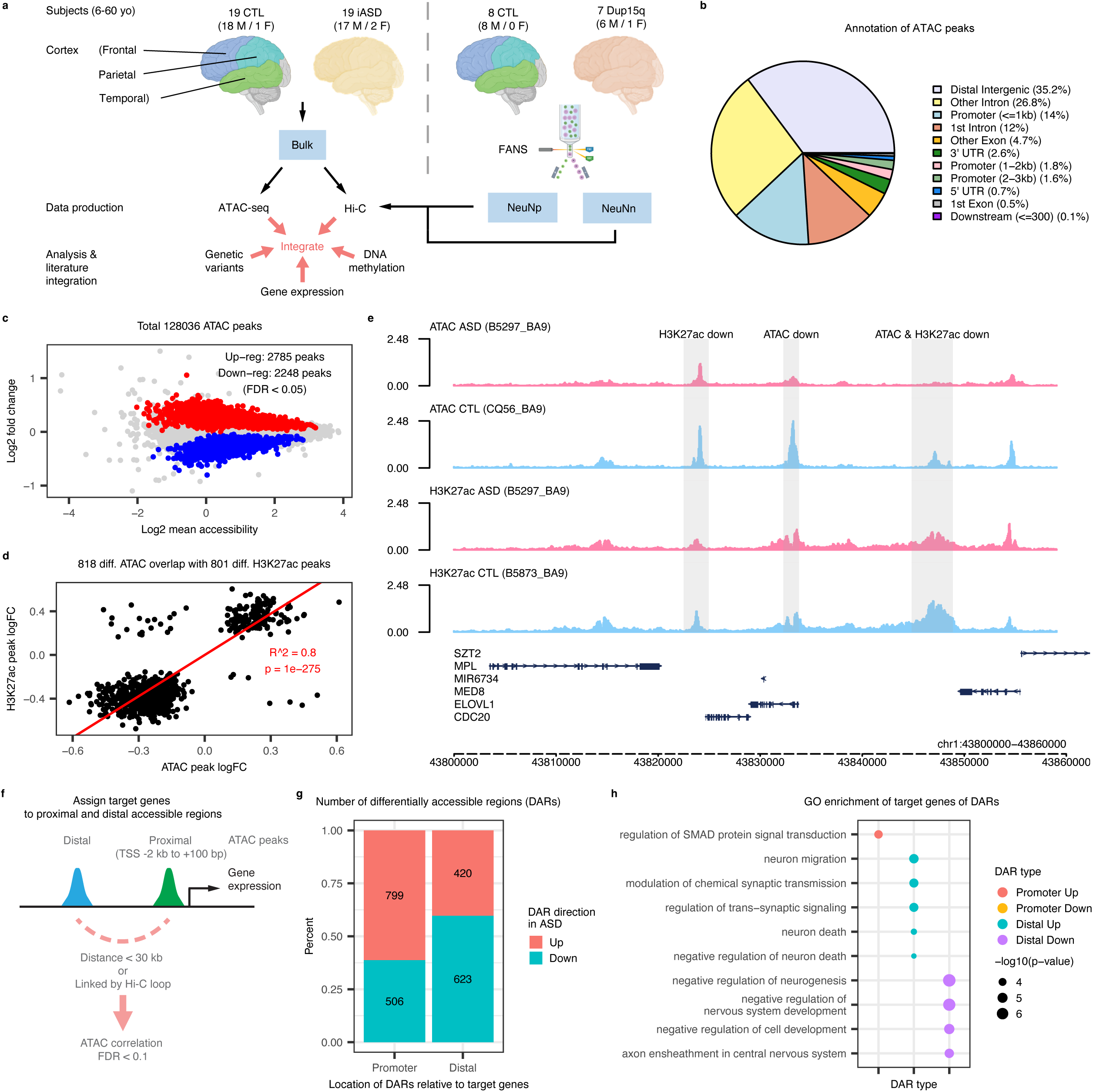
Genome-wide differential chromatin accessibility in ASD post-mortem cortex. **a,** Schematic representation of the study design. **b,** Distribution of consensus ATAC peaks. **c,** MA plot showing differential chromatin accessibility. Up- and down-regulated ATAC peaks (FDR < 0.05) are highlighted in red and blue, respectively. **d,** Scatterplot showing significant correlation between the log fold changes of differential ATAC and differential H3K27ac signals. **e,** Genome tracks showing the dysregulation of ATAC and H3K27ac peaks at chr1: 43,720,000-43,880,000. Dysregulated ATAC peaks and H3K27ac peaks are highlighted by the grey filled boxes (FDR < 0.05). **f,** Schematic of the strategy to assign target genes for proximal and distal accessible regions. Promoter ATAC peaks are defined as those located between 2 kb upstream and 100 bp downstream of a gene TSS. Distal ATAC peaks are assigned to target genes by positive and significant correlation of proximal-distal ATAC pairs either within 30 kb of distance or linked by Hi-C loops (see Methods for details). **g,** Distribution of up- and down-regulated ATAC peaks that have been assigned to target genes at gene promoters and distal regions. **h,** Top gene ontology (GO) terms enriched (FDR < 0.15) in target genes of dysregulated ATAC peaks. Of note, the SMAD protein term FDR = 0.14; for all other terms FDR < 0.1.

We next aimed to identify differentially accessible regions (DARs) between ASD cases and controls. Using a linear mixed model to account for potential confounders (Methods, Supplementary Figure 2), we identified 2,785 up-regulated and 2,248 down-regulated peaks at an FDR < 0.05 (Fig. 1c, Supplementary Figure 3a). To validate the chromatin accessibility changes, we compared our DAR signal with previously identified differential H3K27ac signals in ASD^21^. We found a high level of consistency between differentially accessible ATAC-seq and H3K27ac peaks (Linear regression, R^2^ = 0.8, p-value < 2×10^-16^; Fig. 1d-e, Supplementary Fig. 3b), despite only 2 samples overlapping between our study and the published H3K27ac study^20,21^. Thus, these ATAC-seq results validate the presence of common chromatin changes across individuals with idiopathic ASD, while refining the locations of both the consistently active and differential regulatory regions.

Based on published annotations^28,30,31^, we were able to discern that the majority of the ASD-associated DARs were either constitutively open in both fetal and postnatal stages (45%) or observed primarily in adult brain (35%) (Supplementary Figure 4a), consistent with expectations based on the ages of our subjects. To study the cell-type specificity of the DARs, we leveraged data from single cell genomics^14^, finding that 80% of the DARs in bulk tissue were active in both neurons and glia (Supplementary Figure 4b). Although fewer than 7% of the DARs in single cell ATAC-seq clusters were identified in bulk tissue, and the changes were attenuated in bulk tissue relative to single cell, the relative ranking of changes was conserved (Supplementary Figure 4c).

To understand the potential consequence of the changes in chromatin accessibility, we next identified their target genes. Experimental evidence strongly suggests that distal regulatory regions physically contact gene promoters via chromatin loops during transcription activation^23,27,32^. In line with this model, we observed significantly higher correlation in ATAC signal between peaks connected via Hi-C derived chromatin loop interactions compared to those that were not connected (Supplementary Figure 3c). Leveraging this knowledge, we developed a simple method to integrate these data, <u>C</u>onsensus for <u>H</u>i-C and <u>A</u>TAC <u>R</u>egulatory <u>T</u>argets (CHART) (Methods, Fig. 1f), which allowed us to identify 23,918 significantly correlated distal-promoter ATAC peak pairs either within 30kb of distance or connected via long-range chromatin loops, assigning 15,420 distal regulatory regions to 8,865 genes (Supplementary Table 2). Changes in chromatin accessibility are expected to affect gene expression^30,33,34^. So, we next analyzed the relationship between differential gene expression and chromatin accessibility, which revealed significant correlation between the two modalities at either promoter or distal regulatory regions, supporting our target gene assignments using CHART (Supplementary Figure 3d-e). We also applied the Activity-by-Contact (ABC) model^35^, which predicts enhancer-gene connections based on Hi-C contact maps and enhancer activity derived from H3K27ac or ATAC-seq profiles, identifying 99,351 distal ATAC-gene connections (Supplementary Table 3). These connections showed significant overlap with our CHART-derived distal ATAC-gene pairs (hypergeometric test, OR = 58.9, p-value < 2×10^-16^; Methods). However, we observed no correlation between changes in gene expression and distal enhancer activity within these 99,351 connections defined by the ABC model. Given that CHART-derived ATAC-gene links were strongly supported by the ABC model and were significantly associated with gene expression, we used the CHART-derived distal ATAC-gene pairs for our subsequent analysis.

We next examined the distribution of up- and down-regulated DARs with respect to their target genes. Notably, approximately 60% of the DARs at gene promoters were up-regulated, whereas approximately 60% of distal enhancer DARs were down-regulated (Fig. 1g). Gene Ontology (GO) enrichment analysis revealed that genes with up-regulated promoter DARs are involved in the regulation of SMAD protein signal transduction, which plays an important role throughout neural development^36,37^, whereas genes with down-regulated distal DARs are involved in regulation of neurogenesis and axon ensheathment (Fig. 1h). Target genes of distal down-regulated DARs showed a modest, but significant, overlap with previously identified genes with down-regulated expression in cerebral cortex from subjects with ASD (hypergeometric test, OR = 1.33, p-value = 9×10^-3^). This links, for the first time, a subset of the down-regulated transcripts in ASD brain with specific changes in their regulatory regions that likely contribute to these expression changes.

### Transcription factors associated with ASD dysregulated regulatory regions

We proceeded to interrogate which changes in chromatin accessibility could be causal and which were more likely to be secondary. Causal changes in chromatin accessible regions might be driven by genetic variation, while secondary changes could result from factors such as dysregulated transcription factor (TF) expression and DNA-binding. For example, increased TF expression might lead to an increase in the number of its footprints and/or footprint depths (Fig. 2a). To investigate this, we first used HOMER^38^ to screen for transcription factor motifs that were enriched in the up- and down-regulated DARs (Methods). HOMER analysis revealed a significant enrichment of binding motifs for CTCF, SOX10, SOX15, SOX4, FOXL1, and FOXG1 in the down-regulated DARs. Conversely, the up-regulated DARs exhibited enrichment in TF motifs such as the FOS and JUN family, BNC2, ATOH7, REL, TFDP1, OLIG1, OLIG2, and NFE2 (Fig. 2b, Supplementary Table 4).

**Fig. 2.**
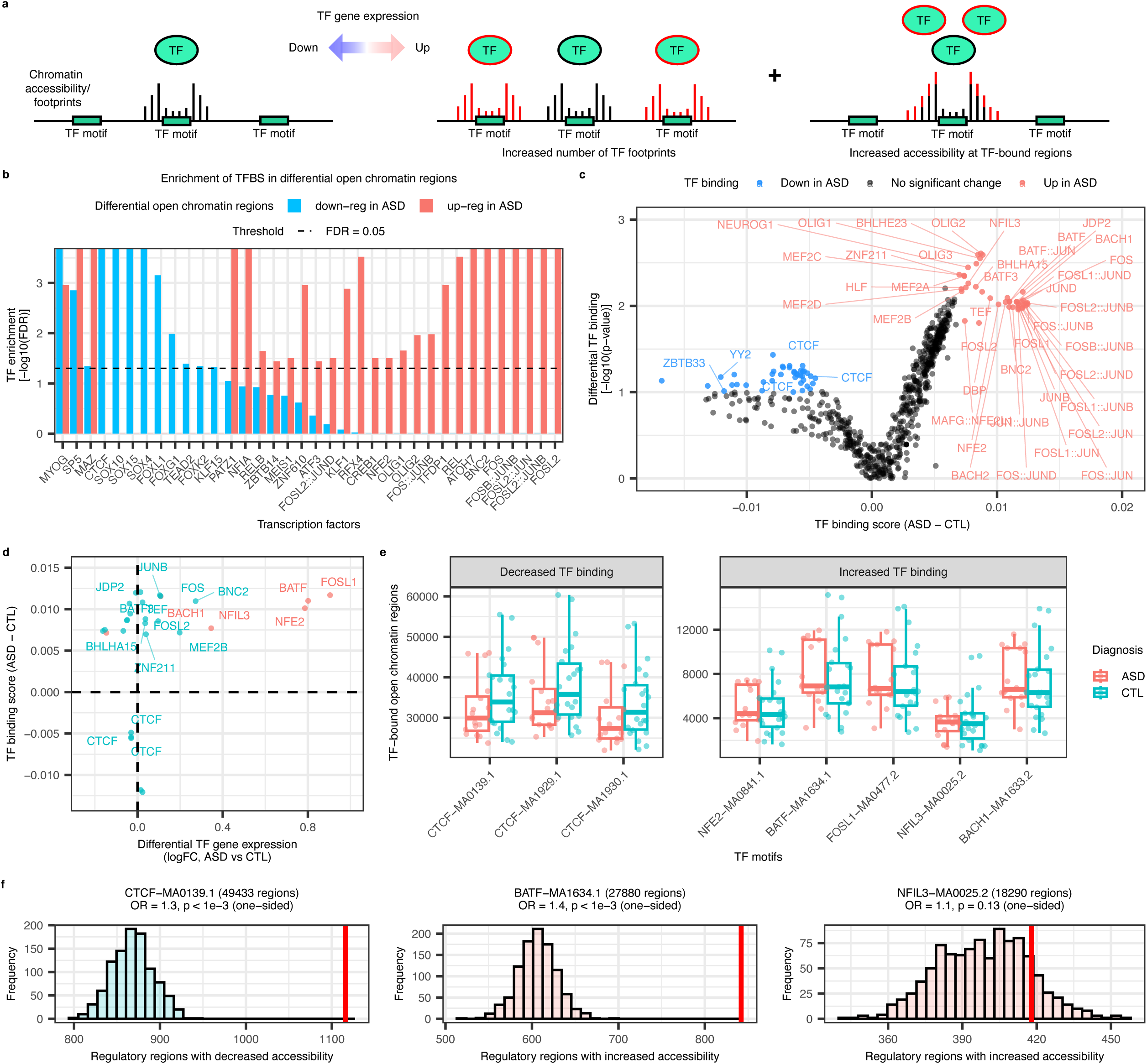
Transcription factors driving dysregulation of chromatin accessibility and transcriptomic changes. **a,** Schematic representation of the hypothesis tested in this figure, that TF gene expression dysregulation influences the accessibility of the TF-bound open chromatin regions. **b,** TF motif enrichment at ASD up- or down-regulated ATAC peaks. **c,** Volcano plot displaying differential TF footprint depth (TF binding score) in ASD vs CTL. **d,** Scatterplot illustrating differential expression of TFs that show differential binding scores. The TFs with significantly dysregulated gene expression are highlighted in red (FDR < 0.05) using gene expression data from Gandal et al., 2022^19^. **e,** Boxplot showing the number of TF-bound ATAC peaks in ASD and CTL samples for the five transcription factors of interest and their binding motifs. **f,** Histograms showing the distribution of the expected number of dysregulated ATAC peaks bound by each TF of interest using a bootstrap strategy. The observed numbers are shown by red bars.

Next, we analyzed TF footprinting using TOBIAS^39^ to validate HOMER predictions and to identify transcription factors with evidence of increased or decreased DNA binding. DNA footprinting analysis validated the decreased DNA-binding of CTCF, as well as increased DNA binding of several of the TFs enriched in the up-regulated DARs in ASD (hypergeometric test, OR = 9.45, p-value = 3×10^-6^; Fig. 2c, Methods, Supplementary Table 4, Supplementary Figure 5a-c). Subsequently, we examined the expression of these TFs in both idiopathic ASD and Dup15q brains^19^ and observed a positive relationship between many of the TF’s footprint depths and expression (Spearman correlation rho = 0.4, p-value = 0.04; Fig. 2d, Supplementary Figure 5d). Among these TFs, *BATF, BACH1, FOSL1, NFE2*, and *NFIL3* showed significantly up-regulated expression in ASD. *CTCF*, which had evidence of reduced binding based on foot-printing, showed significantly down-regulated expression in Dup15q (linear mixed model, FDR = 0.01^19^) and a trend towards down-regulation in ASD versus controls (linear mixed model, FDR = 0.36^19^).

We further interrogated the relationship between TF dysregulation and chromatin accessibility by asking: 1) whether there were changes in the number of the chromatin accessible regions bound by each of these TFs, and 2) whether the accessibility of the TF-bound regions was more likely to change than unbound regions. We found a trend towards a lower number of ATAC peaks bound by CTCF in ASD and a higher number of ATAC peaks bound by the up-regulated TFs (Fig. 2e). Moreover, the chromatin accessibility at ATAC peaks bound by CTCF was significantly more likely to be down-regulated in ASD, while those bound by BATF, BACH1, FOSL1, or NFE2 were significantly more likely to be up-regulated. Although the overall relationship between chromatin accessibility and transcriptional activation is in general known to be modest^40–43^, we observed a significant association between the TF dysregulation and alterations in chromatin accessibility in ASD (OR ranges from 1.2 to 1.4, p-value < 1×10^-3^; Fig. 2f, Fig. 3a-b, and Supplementary Figure 5e). In addition, we found that ATAC peaks bound by the up-regulated TFs were largely shared across all four TFs (Fig. 3c), indicating that these transcription factors may act in combination and bind to the same regulatory regions. To delve deeper, we used DAPPLE and STRING to assess potential physical interactions among these TFs. This affirmed their involvement in a protein-protein interaction (PPI) network (Fig. 3d, Supplementary Figure 5f).

**Fig. 3.**
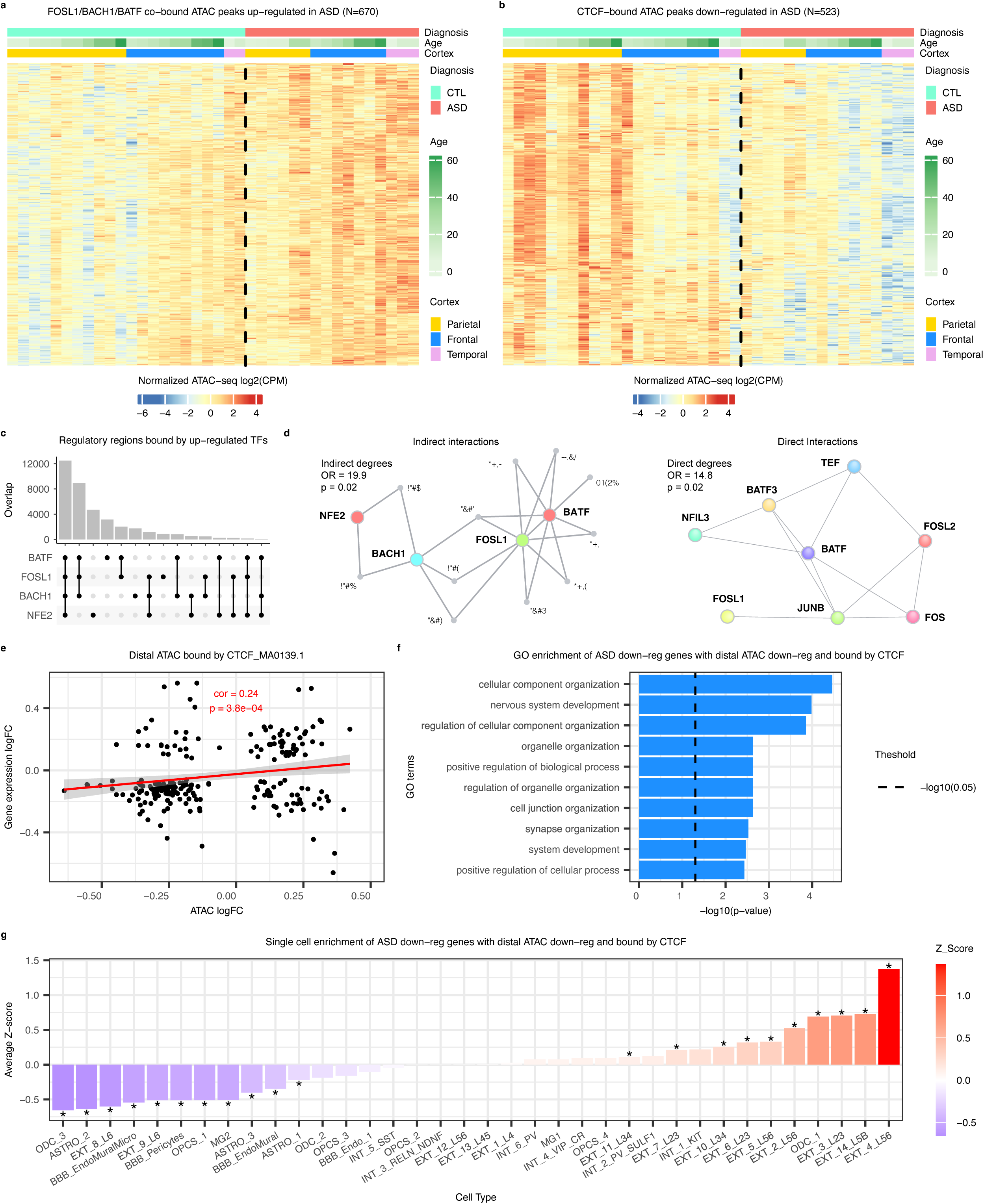
Transcription factor network and target gene function. **a,** Heatmap displaying chromatin accessibility for ASD up-regulated ATAC peaks co-bound by FOSL1, BACH1 and BATF. **b,** Heatmap displaying chromatin accessibility of ASD down-regulated ATAC peaks bound by CTCF. **c,** UpSet plot showing the number of overlapped ATAC peaks bound by BATF, FOSL1, BACH1 and NFE2. **d,** DAPPLE PPI networks showing indirect (left) and direct (right) physical interactions among the up-regulated TFs whose binding scores also increase. **e**, Scatterplot showing significant correlation between the log fold changes in CTCF-bound distal ATAC intensities and target gene expression. **f-g**, Enrichment of GO terms (f) and cell types from Wamsley et al.^14^ scRNA clusters (g) for genes down-regulated in ASD with associated distal down-regulated ATAC peaks bound by CTCF (quadrant 3 of Fig. 3e).

We then tested whether the predicted differential TF binding resulted in differential target gene expression by examining the relationship between target gene expression and chromatin accessibility of the DARs bound by the five dysregulated TFs with changes in DNA footprints (BATF, BACH1, FOSL1, NFE2, and CTCF). For the up-regulated TFs, we observed a significant correlation between the expression of their target genes and corresponding promoter chromatin accessibility changes (Pearson correlation r between 0.25 and 0.41 for various TFs, all p-values < 0.05), but not between the expression of target genes and distal chromatin accessibility changes (Supplementary Figure 6a-b). In contrast, chromatin accessibility at both promoter and distal chromatin accessible regions bound by CTCF showed significant correlations with predicted target gene expression (Pearson r = 0.21 and 0.24 respectively, p-value < 0.05, Supplementary Figure 6c, Fig. 3e). Target genes of BACH1-bound promoters and CTCF-bound distal regulatory regions were supported by the BACH1 and CTCF-regulated gene modules or regulons derived from single-cell RNA-seq data as expected^14^ (Methods, Supplementary Figure 6d-e). The lack of significant overlap between target genes of CTCF-bound promoters and the CTCF regulon suggests that CTCF may contribute to the ASD-associated changes in gene expression mainly via decreased binding to distal regulatory regions. In contrast, a weak correlation between target gene expression and distal ATAC peaks bound by the up-regulated TFs suggests that the up-regulated TFs affect ASD-associated changes mainly via increased binding to promoter regions rather than distal regions.

To investigate functional relevance of the target genes at the single-cell level, we performed Gene Ontology (GO) and cell type enrichment analyses. We found that up-regulated, proximal target genes of the up-regulated TFs significantly overlapped with genes co-expressed in astrocytes and up-regulated in ASD (Supplementary Figure 6f). In contrast, down-regulated, distal target genes of CTCF were enriched in excitatory neurons and oligodendrocytes and pathways involving nervous system development and synapse organization (Fig. 3f-g). Notably, neural cells with the highest number of down-regulated genes in single-nucleus RNA-seq (snRNA-seq) data from ASD brain relative to controls^14^ also ranked among the most affected by decreased CTCF binding at distal open chromatin regions (OR = 12.4, p-value = 0.006; Methods), suggesting that reduced CTCF binding contributes to the observed gene down-regulation in these cell types in ASD. Overall, our findings suggest that the up-regulated chromatin accessibility and gene expression in ASD brains are partially attributed to the up-regulation of the AP-1/NFE2 transcription factors acting in astrocytes. Down-regulated chromatin accessibility and gene expression can be partially attributed to reduced CTCF binding in neurons and oligodendrocytes.

### Common genetic variants at ASD distal down-regulated chromatin accessible regions increase disease risk

We next asked to what extent changes in chromatin accessibility could be explained by genetic risk variants (Methods). To address this, we filtered the catalog of brain chromatin accessibility quantitative trait locus (caQTLs)^28^ to those that were associated with ATAC peaks that overlapped our DARs (Methods, Supplementary Table 5), observing that caQTLs within ASD down-regulated distal ATAC peaks were more likely to increase the risk of ASD (Fig. 4a, Supplementary Figure 7). A salient example of such a caQTL is *rs12977849*, which is associated with a down-regulated ATAC peak on chr19 and is linked to several target genes via promoter-enhancer chromatin loops (Fig. 4b). Two of the linked genes, *NACC1* and *NFIX*, are known ASD risk genes (SFARI database^13^), while *TRMT1* has been associated with intellectual disability^44^. We validated this regulatory relationship experimentally via CRISPR knock-out of the intergenic accessible region or a region containing the risk SNP, both resulting in down-regulation of *TRMT1* expression in primary human neural progenitor cells (Fig. 4c-d, Supplementary Figure 8a-b, Supplementary Table 6). This example illustrates how common genetic variants associated with ASD can impact chromatin accessibility in regulatory regions, changing the expression of risk genes.

**Fig. 4.**
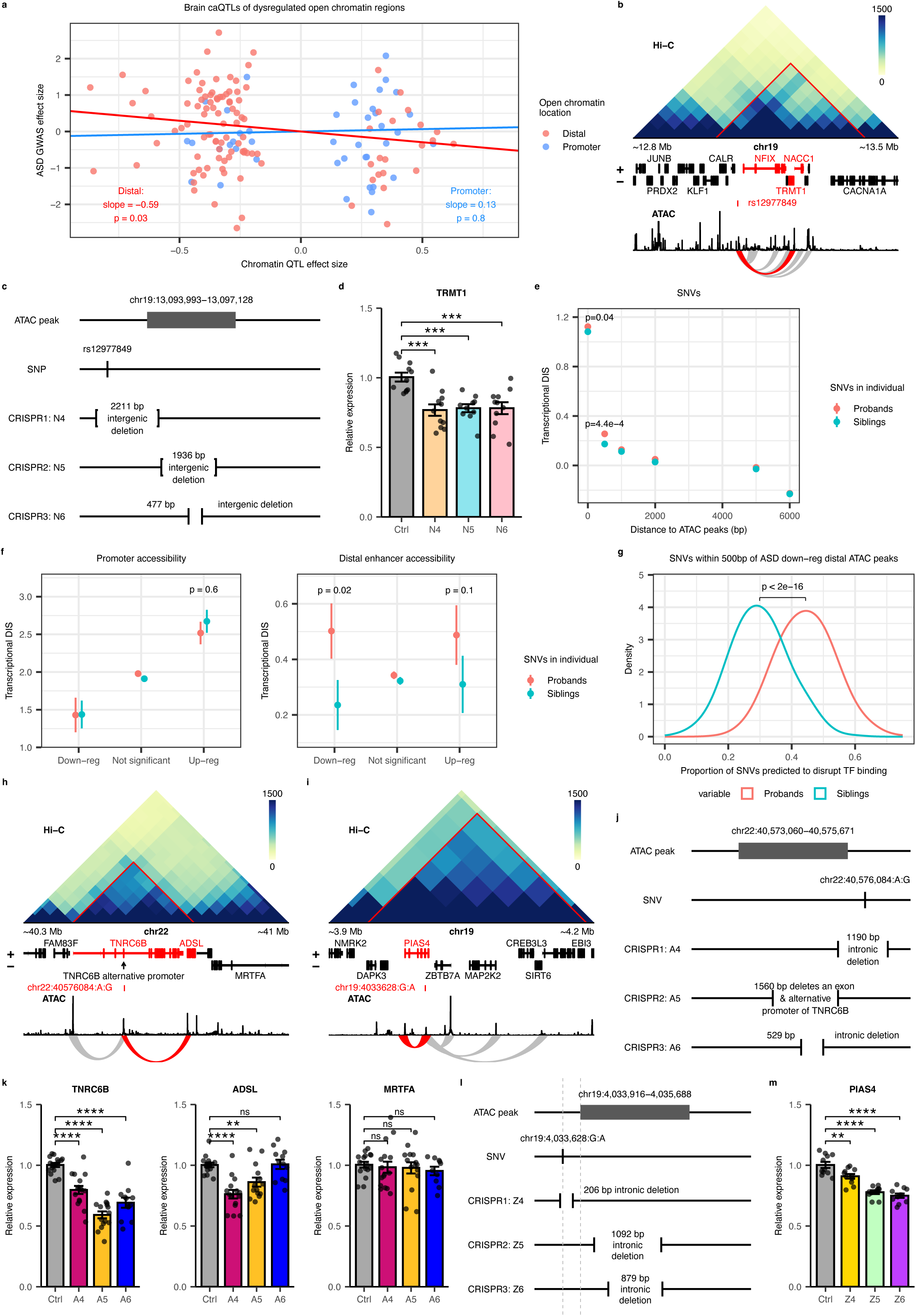
Genetic variants at distal dysregulated open chromatin regions increase ASD disease risk. **a,** Scatterplot showing the correlation between brain caQTL effects on chromatin accessibility at promoter and distal ASD-dysregulated ATAC peaks and their effect on ASD risk. **b,** Hi-C maps and genome browser tracks illustrating the landscape surrounding the brain caQTL (*rs12977849*) at a distal down-regulated ATAC peak. Chromatin loops between the caQTL and promoter regions with significant ATAC signal correlations are highlighted in red, while all other chromatin loops that involve this caQTL are shown in grey. Genes corresponding to those promoters are highlighted in red. **c,** Schematic representation of locations of the targeted ATAC peak, SNP, and three CRISPR/Cas9-mediated genomic deletions at the locus shown in 4b. **d,** qRT-PCR results showing down-regulated expression of the predicted target gene *TRMT1* following CRISPR/Cas9-mediated deletion. **e,** Non-coding *de novo* single nucleotide variants (SNVs) identified in ASD probands versus unaffected siblings show higher transcriptional disease impact score (DIS). **f,** Relationship between DIS and enhancer accessibility highlights distal down-regulated enhancers. **g,** SNVs identified in ASD probands near ATAC peaks disrupt transcription factor binding significantly more than those identified in the unaffected siblings. **h,** Hi-C maps and genomic tracks showing the genomic landscape around the SNV chr22:40576084:A:G identified in ASD proband. The chromatin loop between the SNV and the promoter of SFARI gene *ADSL*, showing significant correlation in ATAC signals, is highlighted in red. Other chromatin loops involving this SNV without significant ATAC correlation are shown in grey. **i,** Hi-C maps and genomic tracks showing the genomic landscape around the SNV chr19:4033628:G:A identified in ASD proband. The chromatin loop between the SNV and the promoter of *PIAS4* with significant correlation in ATAC signals, highlighted in red. Other chromatin loops involving this SNV without significant ATAC correlation are shown in grey. **j,** Schematic representation of locations of the targeted ATAC peak, SNV, and the three CRISPR/Cas9-mediated genomic deletions at the locus shown in Fig. 4i. **k,** qRT-PCR results showing down-regulated expression of the predicted target gene *PIAS4* following CRISPR/Cas9-mediated deletion. **l,** Schematic of the targeted ATAC peak, SNV, and three CRISPR/Cas9-mediated genomic deletions at the locus shown in Fig. 4j. **m,** qRT-PCR showing expression of the two predicted target genes (*ADSL* and *TNRC6B*) and a gene predicted to be unaffected (*MRTFA*) following CRISPR/Cas9-mediated deletion. ** denotes Dunnett-corrected p-value < 0.01, *** denotes Dunnett-corrected p-value < 0.001, **** denotes Dunnett-corrected p-value < 0.0001, while “ns” denotes Dunnett-corrected p-value > 0.05.

### Rare SNVs within DARs impact chromatin structure and transcription

Identification of association signals from rare, non-coding variation has proven difficult^45^, but has been greatly facilitated through epigenome annotations^46–49^. Since common genetic variants in regulatory regions with reduced chromatin accessibility in ASD increased the risk of ASD, we reasoned that we could use these regions to interrogate the impact of rare, non-coding variation. We leveraged the rare *de novo*, non-coding variants identified by Zhou et al^50^ from whole genome sequencing conducted on nearly 2,000 ASD simplex families in the SFARI cohort, including ASD probands and their unaffected siblings. Zhou et al assigned disease impact scores utilizing a deep-learning-based framework to predict the transcriptional and post-transcriptional impact of each variant^50^. Remarkably, our analysis revealed that rare SNVs within 500bp of consensus ATAC peaks in the brain, including the subset of rare SNVs inside the ATAC peaks themselves, exhibited a larger effect on transcription in probands compared to unaffected siblings (Fig. 4e). This relationship is consistent with the GTEx consortium’s observations demonstrating a relationship of rare variation to nearby gene expression in control tissues^51^. When we focused on DARs and separated the ATAC regions based on their direction of change and relative location to gene TSSs, we found that the difference between ASD probands and unaffected siblings could be explained by SNVs within or near the ASD down-regulated distal ATAC regions (Fig. 4f; Methods). Rare variants that disrupt TF binding are expected to have the largest impact on transcription^51–53^, and TF binding information was incorporated into the deep-learning framework used to predict the transcriptional impact of the SNVs in Zhou et al.^50^. Indeed, we found that a significantly higher proportion (OR = 1.5, p < 2e-16) of SNVs in the proband were predicted to disrupt TF binding compared to those in unaffected siblings (Fig. 4g). Two examples of such SNVs are illustrated in Figure 4h-i. One is an A to G mutation at chr22:40576084, located within 500bp of an ASD down-regulated ATAC peak, which we link by CHART to known ASD risk genes, *ADSL* and *TNRC6B* (Fig. 4h). This SNV is predicted to disrupt Sox4 and Sox10 binding motifs (Supplementary Figure 8c). Removing either the ATAC peak or a region containing the SNV resulted in significant down-regulation of both *TNRC6B* and *ADSL* expression (Fig. 4j-k). The other example is a G to A mutation at chr19:4033628, which is linked to multiple genes via chromatin loop interactions and is predicted to disrupt the ZNF135 binding motif (Supplementary Figure 8d). However, only *PIAS4* shows a significant correlation based on the ATAC signal with the SNV-associated open chromatin region, nominating it as the likely target gene (Fig. 4i, Supplementary Table 6). We again validated the regulatory relationships using CRISPR knock-out assays in primary human neural progenitors. When we removed either a small intronic region encompassing the SNV or the ATAC peak located 300bp from the SNV, expression of *PIAS4* showed significant down-regulation, whereas other genes at the locus were unaffected (Fig. 4l-m, Supplementary Figure 8e-f). This supports our target gene prediction and suggests a role for *PIAS4*, which encodes a SUMO E3 ligase, in autism pathogenesis. Taken together, our analyses suggest that common and rare genetic variants contribute to ASD through disruption of ASD-associated distal down-regulated accessible regions.

### Genome-wide changes in the strength of TAD boundaries affect gene expression in ASD

*CTCF* is a known ASD risk gene^54^ whose expression and genome-wide DNA binding were reduced in ASD and Dup15q samples. We hypothesized that this would lead to 3D chromosome structure changes because *CTCF* is known to play a pivotal role in 3D genome organization^23,55–58^. To assess genome-wide changes in 3D genome architecture, we performed Hi-C in ASD and controls, as well as on FANS-sorted neuronal and glial samples from Dup15q brain tissue, aiming to uncover potentially shared but previously uncharacterized cell-type specific chromosome conformational changes (Fig. 1a, Supplementary Figure 9 a-b). The Hi-C data were of high quality (Supplementary Figure 9c, Supplementary Table 7) and revealed the expected 3D genome dynamics between neurons and glia (Fig. 5a, Supplementary Figure 9d-e, Supplementary Figure 10-11, Supplementary Note 1, Methods). Notably, 3D genomic organizational changes were shared between idiopathic ASD and Dup15q syndrome and correlated with chromatin accessibility changes (Supplementary Figure 12-14, Supplementary Note 1, Methods).

**Fig. 5.**
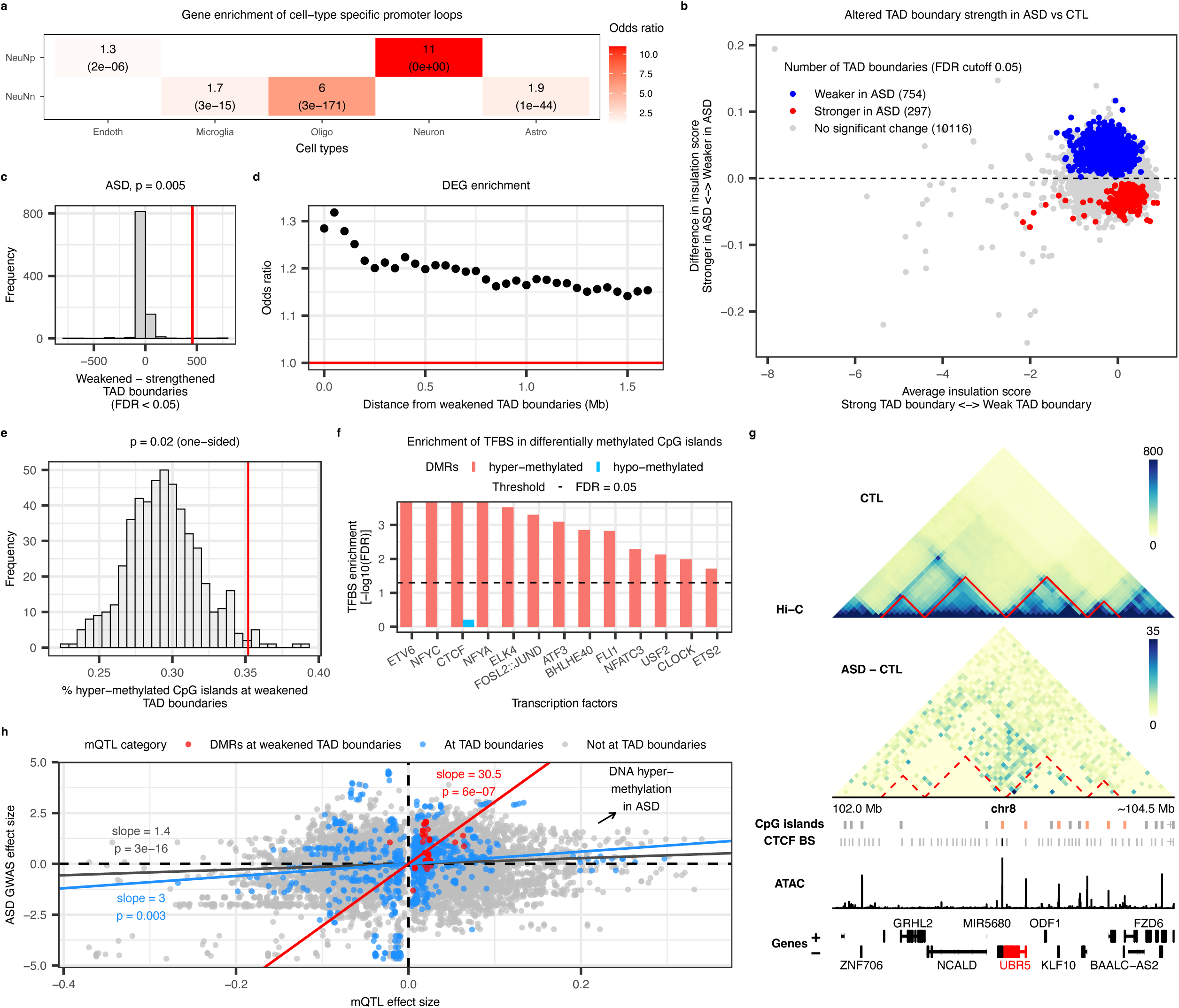
Genome-wide changes in FIREs and TAD boundary strength in ASD subjects. **a,** Enrichment analysis showing that genes at the cell-type specific promoter loops are significantly enriched for the corresponding cell-type specifically expressed genes. Enrichment odds ratios with FDR < 0.1 are labeled. **b,** MA plot showing differential insulation score at TAD boundaries in ASD. TAD boundaries (at 40kb resolution) with reduced or increased insulation (FDR < 0.05) are highlighted in blue and red respectively. **c,** Histograms showing the distribution of the expected difference in the number of weakened and strengthened TAD boundaries in ASD bulk tissue using a permutation strategy. The observed number is depicted (red bar). **d,** Enrichment of differentially expressed genes within 1.5Mb of weakened TAD boundaries. **e,** Histogram showing the distribution of the expected proportion of hyper-methylated CpG islands at TAD boundaries using a bootstrap strategy. The observed proportion at weakened TAD boundaries is shown (red bar). **f,** Enrichment of TF binding motifs in hyper- and hypo-methylated CpG islands (FDR < 0.05). **g,** Genomic landscape changes at the *UBR5* locus. TADs are marked by red triangles on the Hi-C maps. The arrows point to the ASD weakened TAD boundary where there are increased chromatin interactions across the boundary. Hyper-methylated CpG islands are highlighted (red), while all other probed CpG islands are marked in grey. CTCF binding sites at the weakened TAD boundary are highlighted in black, while all other CTCF binding sites are marked in grey. **h,** Scatterplot showing significant correlation between brain mQTLs and their effect on ASD disease risk. The red, blue and grey lines are regression lines for mQTLs associated with ASD differentially methylated regions at weakened TAD boundaries, mQTLs located at all TAD boundaries and those that are not.

We focused on topologically associating domains (TADs), as CTCF binding is enriched at the boundaries and knockdown of CTCF is known to weaken TAD boundaries^59–62^. Consistent with decreased CTCF expression and DNA binding in ASD, we identified 754 weakened and 297 strengthened TAD boundaries in idiopathic ASD (FDR < 0.05, Linear Mixed Model, Methods, Fig. 5b), with the number of weakened TAD boundaries significantly exceeding random expectations (permutation p-value = 0.005, Fig. 5c, Methods). Subjects with Dup15q showed similar genome-wide pattern of weakened TAD boundaries observed in idiopathic ASD (Supplementary Figure 12, p-values <= 0.015). Next, we explored whether changes in TAD boundary strength were correlated between idiopathic ASD and Dup15q by analyzing the log fold changes in insulation scores, observing significant positive associations (Supplementary Figure 14a, Pearson r = 0.13-0.34, p-values < 2×10^-16^).

TAD boundaries are crucial for insulating enhancers from potentially unrelated target genes^57,63^, and it has been reported that expression of genes near TAD boundaries is particularly sensitive to disruptions in TAD formation^62^. So, we next investigated whether DEGs were enriched within 40kb of weakened TAD boundaries, revealing significant enrichment in both idiopathic ASD and Dup15q (Fisher’s Exact Test; ASD: p-value = 5×10^-6^, OR = 1.5; Dup15q neurons: p-value = 1×10^-3^, OR = 1.4; Dup15q glia: p-value = 7×10^-3^, OR =1.8). This enrichment did not depend on choice of an arbitrary distance, as the DEG enrichment extended in a declining gradient up to 1.5 Mb from weakened TAD boundaries (Fig. 5d, Supplementary Figure 15). The direction of gene expression changes (up- or down-regulation) did not show a consistent pattern (Fisher’s exact test, p-value > 0.2), suggesting local genomic context-dependent effects. Importantly, we observed no DEG enrichment near strengthened TAD boundaries (Supplementary Figure 15), consistent with a pathogenic role for the weakening of TAD boundaries in ASD.

### Reduced TAD boundary strength is associated with DNA hypermethylation

The previous observation that DNA hypermethylation at CTCF sites inhibits CTCF binding^64,65^ prompted us to test the hypothesis that CTCF binding sites at the weakened TAD boundaries might be hypermethylated. Re-analyzing DNA methylation data from 42 ASD and 27 control brains^66^ (Methods), we identified 8907 differentially methylated regions (DMRs), with 6638 displaying hypermethylation and 2261 showing hypomethylation in ASD (Supplementary Table 8). There was a significant enrichment of hypermethylated CpG islands at the weakened TAD boundaries in ASD (permutation p-value = 0.02, Fig. 5e, Methods). Across hypermethylated regions genome-wide, TF binding site enrichment analysis (Methods) highlighted CTCF among the enriched TFs in the hypermethylated regions (Fig. 5f), whereas no enrichment of any TF was observed at hypomethylated regions. We illustrate the association between hypermethylation of the CTCF binding site and weakened TAD boundaries at the *UBR5* locus (Fig. 5g), which codes for a high-confidence ASD risk gene (SFARI database^13^) located within 40kb of a weakened TAD boundary and is significantly up-regulated in ASD in post-mortem brain^19^. We observed hypermethylation of the CpG island encompassing a CTCF binding site at this TAD boundary.

We next sought to understand whether ASD genetic risk might act through its impact on DNA methylation and TAD boundaries by examining the relationship between the effect size of mQTLs^67^ on DNA methylation and their effect size on ASD risk, as ascertained by GWAS (Grove 2019). This analysis revealed that mQTLs that are predicted to increase DNA methylation were more likely to increase the risk of ASD, particularly when they were located within DMRs at the weakened TAD boundaries (Fig. 5h, Methods; linear regression, beta = 30.5, p-value = 6×10^-7^). In summary, our investigation suggests a causal association between genetic risk for ASD, DNA hypermethylation at CTCF binding sites, and weakening of TAD boundaries.

### Individual contributions and interplay among multiple ASD-associated molecular profiles

Having revealed alterations in chromatin accessibility at both promoter and distal regulatory regions, along with weakened TAD boundary strength in ASD, and demonstrated their connections with dysregulated gene expression (Fig. 6a), we examined the individual contributions of each of these factors to the entire group of DEGs observed previously in brain^19^ (Fig. 6b-c, Methods). Among the up-regulated DEGs with increased chromatin accessibility at their promoter regions, 33% were bound by the up-regulated AP-1 and NFE2 transcription factors. In contrast, among the down-regulated DEGs exhibiting decreased chromatin accessibility at the promoter and distal regulatory regions, CTCF binding was observed in 91% of cases. Notably, we showed that distal regulatory regions manifesting decreased chromatin accessibility were enriched in common ASD risk variants. There was little overlap between the DEGs affected by chromatin accessibility changes and those within 40kb of weakened TAD boundaries, suggesting an independent effect of CTCF binding on TAD boundaries and regions with changes in chromatin accessibility. Collectively, these changes could explain 20% of all DEGs observed in ASD brain (Fig. 6c, Supplementary Table 9).

**Fig. 6.**
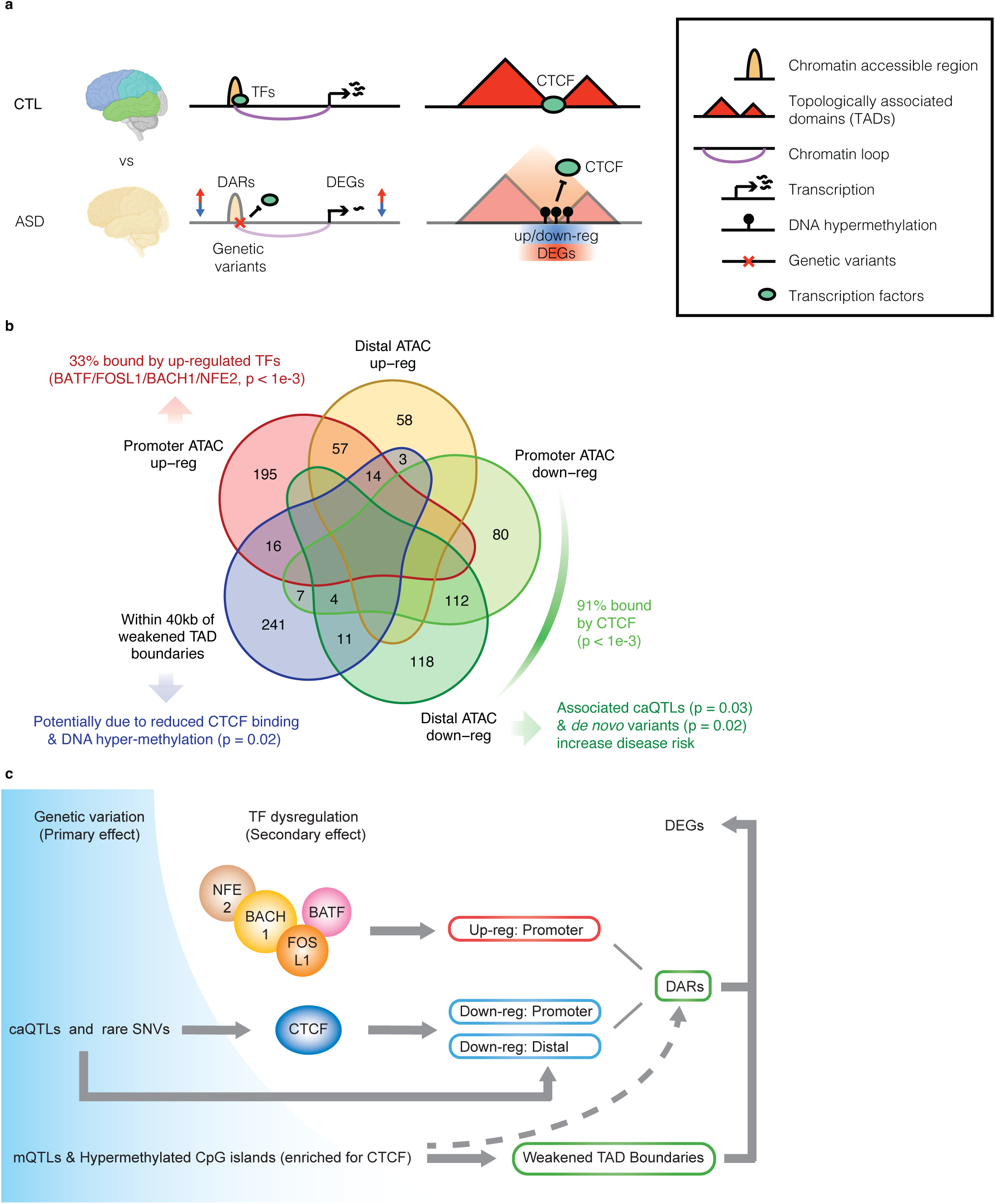
A working model for ASD etiology. **a,** Schematic of the observed changes in chromatin accessibility and strength of TAD boundaries in ASD brains, and their impact on gene expression. **b,** Schematic representation of the contribution of the five dysregulated epigenetic regions to ASD-associated differentially expressed genes (DEGs). The number of DEGs associated with each epigenetic region is labeled. The connections between changes in transcription factor expression and footprints, DNA methylation, and genetic variants with the dysregulated regions are indicated by arrows. **c,** Schematic representation of the genetic mechanism of ASD. The percentage of DEGs attributed to each genetic and epigenetic factor is indicated in parentheses.

## Discussion

Through extensive analysis of chromatin architecture in human brain, we identify genome-wide changes in regulatory architecture that provide a new view of the genetic mechanisms and subsequent transcriptional dysregulation in ASD brain. More than a decade of study has revealed consistent changes in gene expression that define the molecular state of the brain in neuropsychiatric disorders. Yet, the mechanisms underlying these changes and to what extent they are driven by known genetic risk factors are not known. We find that several different epigenetic alterations, most prominently changes in chromatin accessibility and weakening of TAD boundaries, coupled with decreased CTCF binding and DNA hypermethylation, are associated with the down-regulation of neuronal and synaptic genes in ASD. These changes in chromatin accessibility and TAD boundaries are associated with brain caQTLs and mQTLs that increase ASD disease risk, consistent with a causal relationship whereby these common elements of ASD genetic risk act via influencing local chromatin accessibility or DNA methylation, both of which influence downstream gene expression. Similarly, the decreased accessibility observed between ASD probands and controls can be explained by rare non-coding SNV within close proximity, or in distal dysregulatory open chromatin regions. We use CRISPR-based gene editing to validate the predicted gene regulatory impact at several of these common and rare variant loci. These findings illustrate how non-coding variants may affect disease risk and brain gene expression by altering distal enhancer accessibility and the 3D genome organization, in both idiopathic and syndromic ASD.

The identification of non-coding variants associated with ASD has been challenging, primarily because their effects are predicted to be smaller compared with protein disrupting variants^50,68^. Additionally, ASD is a complex neurodevelopmental disorder involving many risk genes^6,69^. Due to these challenges, studies attempting to identify non-coding variants associated with ASD have been limited by power^70–73^. Notably, no single non-coding annotation category has consistently passed the threshold of genome-wide significance in ASD association, despite the prominence of promoter regions in some studies^7,73^. Leveraging our differential chromatin accessibility data, we found that both common and rare variants located within 500bp of the distal down-regulated accessible regions contribute to increased ASD risk. Moreover, mQTLs associated with elevated DNA methylation, particularly those within DMRs at weakened TAD boundaries, exhibit a significant effect on increased ASD risk. This finding aligns with prior research indicating that DNA hypomethylation at specific CTCF sites can lead to dysregulation of chromatin accessibility and medical conditions typically related to mutations in risk genes, for example, at the *FTO* locus in BPA-induced obesity^74^. Additionally, transcriptomic signatures themselves have been demonstrated to increase power to identify single nucleotide and structural variants with functional impact^75–77^. These analyses and others^77–80^ support the value of incorporating disease-relevant, differential epigenomic and transcriptomic findings into the process of genetic variant prioritization and identification.

CTCF is a transcription factor that has multi-faceted effects on gene regulation, at the promotor via direct DNA binding, and at distal regulatory regions by shaping 3D chromatin structure, including enhancer-promotor interactions via its anchoring effects on chromatin loops. *CTCF* mutations cause a range of developmental disorders in which ID/DD and language delays are prominent and is also an ASD risk gene^54^. Down-regulation of *CTCF* expression and its DNA footprints genome-wide, combined with DNA hypermethylation, are predicted to contribute to the substantial decrease in the strength of TAD boundaries. Although with these data we are unable to directly establish the temporal sequence linking dysregulated DNA methylation and *CTCF* expression and binding, the alterations in chromatin accessibility, chromosome conformation, and transcriptional dysregulation serve as promising targets for therapeutic intervention and underscore the importance of investigating epigenetic factors in understanding ASD.

Several limitations of this study are worth highlighting. The ability to understand the contributions of genetic risk factors to the changes in brain is limited by the relatively small percentage (<2-3%) of risk variance explainable by published GWAS in ASD^81^. In this context, the significant relationship between known genetic risk and changes in chromatin and methylation that we observe is remarkable. Additionally, while we anticipated detecting widespread alterations in chromatin loop interactions that would directly explain changes in gene expression, our findings emphasize the need for even larger datasets to capture more subtle but significant alterations. Furthermore, both our Hi-C and ATAC-seq analyses underscore the importance of cell-type specific profiling, as between 25 and 40% of 3D chromatin loops were uniquely found in each cell type (Fig. 4d).

Taken together, our analyses provide a framework that integrates multiple functional genomic analyses to understand how changes in DNA sequence, DNA methylation, chromatin accessibility, and chromosome conformation may interact to contribute to molecular changes in brain. We linked distal regulatory regions to target genes based on correlation of chromatin accessibility between promoter and distal regions, coupled with their 3D chromatin interactions. We further analyzed transcription factor footprints to determine their binding at specific sequences and link to chromatin accessibility and gene expression changes. We inferred causal relationships by assessing the disease risk of the genetic variants in sub-categories of chromatin accessibility regions and differentially methylated regions, highlighting critical roles of genetic variants at distal regulatory regions and insulation boundaries. Moving forward, it would be valuable to interrogate these ASD-associated changes in a cell-type specific manner across neurodevelopment, which is limited by current technology.

## Data Availability

All data produced in the present study are available upon reasonable request to the authors

## Acknowledgements

Funding for this work was provided by grants to D.H.G. (NIMH R01MH110927) and J.Y. (Training Grant in Neurobehavioral Genetics, Award Number T32NS048004). We thank the UCLA Broad Cell Research Center Flow Cytometry Core and Sequencing Core, as well as the UCLA Technology Center for Genomics & Bioinformatics for their expertise. Additionally, we would like to thank Drs. Yan Jiang and Schahram Akbarian from Mount Sinai School of Medicine for sharing their nuclei isolation protocol.

## Methods

### Sample acquisition and dissection of post-mortem brains

Post-mortem cortical brain samples were acquired from the Autism BrainNet project (ABN), formerly the Autism Tissue Project at Harvard (ATP) and the University of Maryland Brain and Tissue Bank (UMDB/NICHD-BTB) through the NIH NeuroBioBank. A total of 53 samples from individuals with ASD or Dup15q syndrome, and neurotypical controls (48 unique subjects) in either of the three cortical regions – frontal: BA4/6, BA9, BA44/45, BA24; parietal: BA3/1/2/5, BA7, BA39/40; and temporal: BA38 – were acquired (Supplementary Table 1). Among the cortical samples used in this study, 39 samples from 35 unique subjects were previously subjected to bulk transcriptomic profiling^16,19^. 2 cortical samples from 2 unique subjects were previously subjected to bulk H3K27ac profiling^20,21^. 14 subjects had not been previously profiled.

Frozen brain samples were securely stored in individual sealed plastic bags at -80 °C. During the dissection process, cortical samples were placed onto a pre-cooled platform in a chamber filled with dry ice to assure preservation of chromatin structure. 100-210 mg of cortical tissue was dissected and used for generating bulk ATAC-seq and Hi-C libraries for the ASD and their matched control samples, and 110-910 mg for nuclei isolation and sorting prior to generation of cell-type specific Hi-C libraries for Dup15q and their matched control samples.

### Nuclei isolation and sorting

To generate bulk and cell-type specific Hi-C libraries, nuclei were extracted from human post-mortem cortical samples as described before^82^. Briefly, aliquots of 100-300 mg of cortical samples were homogenized by douncing in an ice-cold hypotonic lysis solution (0.32M Sucrose; 5mM CaCl_2_; 3mM Mg(Ace)_2_; 0.1mM EDTA; 10mM Tris-HCl, pH8; 1mM DTT; 0.1% Triton X-100). Brain homogenate was fixed with 1% formaldehyde for 5 min at room temperature, followed by 2M Glycine quench for 5 min and bench-top centrifuge at 4000 rpm. The resulting pellet was rinsed with the lysis solution and filtered through a 100µm cell strainer prior to ultracentrifuge at 100,000 g at 4°C for 1 hour in a sucrose gradient – top layer: 5ml of the nuclei solution; bottom layer: 9ml of sucrose solution (1.8M Sucrose; 3mM Mg(Ace)_2_; 10mM Tris-HCl, pH8; 1mM DTT). For the ASD and their matched control samples, the nuclei pellet was stored at -80°C. To immuno-label the neuronal proportion in the Dup15q and their matched control samples, the nuclei pellet was resuspended in 1ml DPBS containing 0.1% BSA and 1:1000 AlexaFluor 488-conjugated antibody against neuronal marker NeuN (MAB377X, Millipore). Samples were incubated with rotation in the dark at 4 °C for at least 45 min. DAPI (62248, Thermo Fisher Scientific) was added before FACS to label all the nuclei. NeuN-positive (NeuNp) and NeuN-negative (NeuNn) nuclei were sorted into separate collection tubes pre-coated with 10% BSA at the Flow Cytometry Core at UCLA, recovered by centrifuge at 2,500 g at 4°C for 5 min, and stored at -80°C. On average, we obtained a ratio of 1:3 between NeuNp and NeuNn nuclei.

### ATAC-seq library preparation and sequencing

ATAC-seq was performed based on the Omni-ATAC protocol as previously described^25^ with minor modifications. Briefly, approximately 40mg of cortical sample was homogenized by douncing (roughly 40 times) in 2ml of ice-cold homogenization buffer (5mM CaCl_2_; 3mM Mg(Ac)_2_; 4mM Tris, pH 7.5; 6mM Tris, pH 8; 0.1mM PMSF), followed by centrifuge at 100 g at 4°C for 1 min. The supernatant was mixed with equal volume of 50% iodixanol prior to centrifuge at 3,000 g at 4°C for 30 min in an iodixanol gradient – top layer: 4ml of sample; middle layer: 3ml of 29% iodixanol; bottom layer: 3ml of 35% iodixanol. 100,000 nuclei from the resulting pellet were transferred into 1ml of resuspension buffer (RSB, 10mM Tris-HCl, pH 7.5; 10mM NaCl, 3mM MgCl_2_; 0.1% Tween-20), centrifuged at 500 g at 4°C for 10 min, and rinsed with 1ml RSB. Pelleted nuclei were resuspended gently in 100µl transposition mix (50µl 2x TD Buffer; 5µl TDE1 transposase from Nextera DNA library prep kit – final concentration 100nM; 33µl DPBS, 1µl 1% Digitonin, 1µl 10% Tween-20, 10µl H2O) and split evenly into two reaction tubes, with each tube serving as a technical replicate for the ATAC-seq library. The mixture was incubated at 37°C for 30 min in a thermomixer. The DNA was purified using the Zymo DNA Clean and Concentrator-5 Kit (D4014), followed by PCR amplification of 3 cycles in a 50µl reaction with Illumina Nextera adaptors and NEBNext Master Mix. To determine the required number of additional cycles for library amplification, a side qPCR reaction was performed using 5µl of the pre-amplified DNA. We used the Ct value of the qPCR at ¼ maximum fluorescence as the number of additional cycles. After running the additional PCR cycles on the remainder of the pre-amplified DNA, the DNA product was purified using the Zymo DNA Clean and Concentrator-5 Kit. The DNA concentration was quantified by the KAPA Library Quantification Kit prior to library pooling and sequencing to an average depth of 83M reads per sample on the Illumina NovaSeq platform at 2×100 bp.

## ATAC-seq data processing

### Read alignment and peak calling

Raw sequencing reads were assessed using FastQC^83^ (version 0.11.3, http://www.bioinformatics.babraham.ac.uk/projects/fastqc) to verify that the general read quality was sufficient prior to further processing following the guidelines provided by HarvardFAS Informatics (https://informatics.fas.harvard.edu/atac-seq-guidelines.html). Paired-end sequencing reads were processed through NGmerge^84^ (version 0.3, https://github.com/jsh58/NGmerge) to remove adaptors and low-quality reads. The processed reads were aligned to the human hg19 reference genome using bowtie2^85^ (version 2.2.9, http://bowtie-bio.sourceforge.net/bowtie2) using--very-sensitive mode, and sorted using Samtools^86^ (version 1.9). Mitochondria reads were removed using removeChrom.py (https://github.com/harvardinformatics/ATAC-seq/blob/master/atacseq/), followed by duplicate removal using Picard Tools (http://broadinstitute.github.io/picard), and the resulting bam files of uniquely aligned non-mitochondria reads were converted to bed files using SAMtoBED.py (https://github.com/harvardinformatics/ATAC-seq/blob/master/atacseq/). MACS2^87^ (version 2.1.2) was used to call peaks on each ATAC-seq library: macs2 callpeak -t [input bed file] -f BEDPE -n [prefix of output file name] -g hs --keep-dup all --nomodel --shift -100 --ext 200. Peaks that overlapped with ENCODE hg19 blacklisted regions were removed.

### Quality control of ATAC-seq libraries

Each ATAC-seq library was subject to the following 5 quality control (QC) criteria: 1) Enrichment of ATAC reads at the TSSs, an important feature of accessible regions, was measured using tssenrich (https://github.com/anthony-aylward/tssenrich). A minimum TSS enrichment score of 3 was required; 2) Read statistics including the insert size was calculated using the Picard Tools CollectInsertSizeMetrics function. Insert or fragment size distribution was assessed based on the guidelines provided in Ou et al 2018^88^, and sub-optimal or over-transposed ATAC-seq libraries were considered to have failed QC; 3) Fraction of reads in peaks (FRiP) was calculated using the bedtools intersect function (version v2.30.0, https://github.com/arq5x/bedtools2), and a minimum FRiP of 0.05 was required; 4) A minimum of 20,000 blacklist-filtered peaks was required; 5) All ATAC-seq libraries satisfied a minimum of 25M uniquely aligned non-mitochondria reads. ATAC-seq libraries that met all 5 requirements were considered successful and included in the final dataset.

We used the DiffBind R package (version 2.14.0, R version 3.6.1) to assess the correlation of ATAC peak signal across the biological and technical replicates: Peaksets <-dba(sampleSheet="SampleSheet.csv"); plot(Peaksets). The minimum correlation between samples was 0.8, and the 6 pairs of technical replicates clustered together.

### Identification of consensus ATAC-seq peaks and peak annotations

To obtain a consensus peak set, technical replicates (ATAC-seq library names: B4334-1, B4721A, B5000B, B5718D, B5813B, CQ56-2) were removed and DiffBind (Stark 2011) was used to calculate the overlap rate of ATAC peaks across the remaining 32 samples. We kept peaks overlapped in at least 5 samples, as this threshold represented the knee point of the overlap rate curve and suggested a high degree of agreement among the peak sets. With this threshold, we obtained a total of 128,036 consensus peaks. The consensus peaks were annotated using ChIPseeker^89^ (version 1.34.1) with gene TSSs derived from the TxDb.Hsapiens.UCSC.hg19.knownGene package (version 3.2.2).

## Differential analysis on chromatin accessibility

We used the featureCounts function from the Subread R package^90^ to obtain the number of reads in the consensus peaks across all samples, including the technical replicates. GC content of each peak was calculated using the bedtools nuc function. We normalized the peak read count matrix using the cqn package^91^ (version 1.44.0), accounting for peak GC content, peak width, and the total number of uniquely aligned non-mitochondrial reads in each library. We then performed principal component analysis (PCA) on the cqn-normalized peak logRPKM matrix using the prcomp function in R, and assessed the correlations of biological and technical covariates with the top 14 PCs which captured around 80% of the total variance (Supplementary Figure 2). We fit linear models for the normalized read counts, incorporating covariates that displayed significant correlation with the top PCs (FDR < 0.2). Colinearity among the covariates was evaluated using the olsrr package (version 0.5.3). The final linear model was chosen by ensuring that all covariates included in the model had a variance inflation factor (VIF) of < 2.5, and we treated technical replicates as a random effect in a linear mixed model: lme(Peak accessibility in logRPKM ∼ Diagnosis + Age + Batch + RegionBA38 + RegionBA44_45 + tssenrich.score + FRiP, random = ∼ 1|Subject). Chromatin accessible regions with an FDR-corrected p-value of < 0.05 between the ASD and control samples were considered significantly dysregulated. The coefficients of the diagnosis term in the linear model for each consensus peak were referred to as the logFC values.

### Overlap and correlation between differential chromatin accessibility and H3K27ac

Differentially acetylated regions were obtained from Ramaswami et al.^21^ and restricted to those with FDR < 0.05. The significance of overlap between differentially accessible regions and differentially acetylated regions in base-pairs was calculated using Fisher’s Exact Test, with the merged regions of consensus ATAC peaks and H3K27ac peaks set as the background for comparison. Pearson’s correlation was calculated between the logFC values of the overlapped differential regions. These results were shown in Fig. 1d-e.

### Quantification of the number of DARs in fetal and adult brains

For adult brain chromatin accessible regions, we used the 118,152 ATAC peaks identified by Bryois et al. in adult prefrontal cortex brain samples from 272 individuals^28^. For fetal brain chromatin accessible regions, we merged the ATAC peaks from two datasets: 1) 62,005 ATAC peaks identified by de la Torre-Ubieta et al. in the germinal zone and cortical plate of post-conception week 15-17 human fetal brain^30^; 2) 64,042 non-promoter open chromatin regions (OCRs) in mid-gestation human prefrontal cortex, parietal and temporal cortex^31^. Constitutive ATAC peaks were defined as those that overlapped by at least 1 bp between the fetal and adult datasets, while the other ATAC peaks were labeled as fetal or adult-specific accessible regions. Promoter regions were defined as 2 kb up- and down-stream of TSSs of all protein-coding genes in the GRCh37.p13 Ensembl database, and the other ATAC peaks were referred to as distal accessible regions. Then we quantified the ASD-associated DARs that overlap these fetal, adult, and constitutive promoter and distal ATAC peaks.

### Differential single-nucleus ATAC-seq analysis

We acquired single-nucleus ATAC-seq (snATAC-seq) data from Wamsley et al.^14^, in which reads were mapped to GRCh38 and data was stored as a Cell Ranger object^92^. We used ArchR^93^ (version 1.0.2) to filter out low quality cells with TSS < 2, minimum number of fragments < 3162 (or 10^3.5). We performed dimensionality reduction using the addIterativeLSI function with the following parameters: resolution = c(0.1, 0.2, 0.4) and varFetures = 15000, and removed batch effect using the addHarmony function. Subsequently, cells were clustered using the addClusters function with resolution = 0.8 and clusters with fewer than 1000 cells were removed, yielding 23 clusters. We annotated these cell clusters based on chromatin activity of marker genes: *SLC17A7* and *SATB2* for excitatory neurons; *LHX6, SST, PVALB, GAD2, SLC32A1*, and *ADARB2* for inhibitory neurons; *SLC1A2, ALDH1A1,* and *GFAP* for astrocytes; *CSPG5, CCND1,* and *OLIG2* for oligodendrocyte progenitor cells (OPCs); *SOX10, MAL, MBP,* and *CLDN11* for oligodendrocytes (ODCs); *IRF8, RUNX1, PTPRC,* and *SLC2A5* for microglia. In addition, we compared the gene activity in snATAC-seq clusters to scRNA-seq clusters from the same samples^14^ using the addGeneIntegrationMatrix function with the top 4000 variable genes on randomly-sampled 40,000 snATAC-seq cells and 25,000 snRNA-seq cells. Multiple snATAC-seq excitatory neuron clusters showed highest similarity to deep layer 6 neurons (EXT_8_L6 in Wamsley et al.) and were merged together, while a distinct excitatory neuron cluster showed highest similarity to superficial layer 2/3 neurons (EXT_6_L23 in Wamsley et al.). We constructed pseudobulk replicates using cells from ASD and CTL individuals separately for each cell cluster and identified reproducible ATAC peaks using the addReproduciblePeakSet function. The ASD and CTL peak sets were merged to generate the final set of reproducible scATAC peaks for each cell cluster and assessed for overlap with DARs from bulk tissue. To identify differential ATAC peaks in each cell cluster, we used the getMarkerFeatures function comparing pseudobulk replicates of ASD and CTL with an FDR cutoff of < 0.05.

### Assign target genes for proximal and distal accessible regions

We acquired the gene TSSs of all 24,836 genes expressed in the human brain cortex (Haney 2022 Nature) using the biomaRt R package (version 2.54.0). Promoters were defined as the regions that spanned 2 kb upstream and 100 bp downstream of the gene TSSs. The consensus ATAC peaks that overlapped with these promoter regions were assigned to the corresponding genes.

To assign target genes for distal accessible regions, we utilized the chromatin loops identified from our Hi-C data. We obtained the union set of consensus chromatin loops in bulk tissue, neurons, and glia at 10 kb resolution, and subset the chromatin loops to those with at least one end overlapping a gene promoter ATAC peak. ATAC peaks that intersect the other end of these promoter loops were referred to as the distal ATAC peaks and paired with the corresponding promoter ATAC peaks. In addition, ATAC peaks located within 30 kb of a gene TSS were paired with that promoter ATAC peak and added to the pool of distal-promoter ATAC pairs. The sample-wise Pearson’s correlation of the cqn-normalized ATAC logRPKM values was calculated for each distal-promoter ATAC pair. Next, to evaluate the significance of the distal-promoter ATAC correlations, we generated a null distribution for each promoter ATAC peak in the pool by randomly sampling 10,000 distal ATAC peaks from a different chromosome and calculating their correlations with the promoter ATAC peak. Distal-promoter ATAC pairs with FDR < 0.1 were used to assign target genes to distal ATAC peaks. Using this approach, we assigned 8,865 genes to 15,420 distal ATAC peaks.

### Link ATAC peaks to target genes using the Activity-by-Contact (ABC) model

We implemented the ABC package^35^ (version 1.0) to predict enhancer-target gene connections, using the corresponding ATAC-seq and Hi-C files of each subject in our study as the input files. The tagAlign files were generated from bam files containing bulk ATAC-seq uniquely mapped reads using the samtools view and bedtools bamtobed functions. The Hi-C allValidPairs files were converted to hic files using the HiC-Pro hicpro2juicebox function. High confidence ATAC-target gene connections were determined by the occurrence with a default threshold of ABC score > 0.02 in at least 1/3 of the subjects (i.e. 10 of the 32 subjects). The resulting 99,351 high confidence ATAC-target gene connections are provided in Supplementary Table 3. To assess the overlap of ATAC -gene connections predicted by the ABC model and those predicted using our home-brewed approach, we performed Fisher’s Exact Test using all possible ATAC peak-gene pairs within 5 Mb of distance as the background connection set.

### Quantification of the number of DARs at gene promoters and distal regions

Differentially accessible regions (DARs) were classified into three categories based on their locations: promoter ATAC, distal ATAC linked to promoters by Hi-C loops, and other distal ATAC peaks. The numbers of up- and down-regulated DARs were quantified and their relative percentages in each category were plotted.

We assigned target genes to DARs that belong to the first two categories using the approach mentioned above. We used the enrichGO function in the clusterProfiler package (version 4.6.2) for GO enrichment analysis of target genes, with the 24,836 brain expressed genes as the background gene list and set the other parameters as following: keyType = “ENSEMBL”, OrgDb = “org.Hs.eg.db”, ont = “BP”. Then we filtered for level 4 or 5 GO terms using the gofilter function. The -log_10_(p-value) of the top GO terms in each DAR category were shown.

### Transcription factor motif enrichment analysis

We used HOMER (version 4.11.1) to identify the transcription factor (TF) motifs that were enriched in the regions of interest (up- and down-regulated ATAC peaks, or hyper-methylated CpG islands) in the ASD brains: findMotifsGenome.pl [bed file] hg19 [output directory] -size given -nomotif -mknown [.motifs file]. We restricted the motif search to transcription factors in the Jaspar 2022 core vertebrates database (JASPAR2022_CORE_vertebrates_non-redundant_pfms_jaspar.txt) that were expressed in the human brain cortex based on Gandal et al.^19^. An FDR cutoff of < 0.05 was utilized to determine the enriched TF motifs.

### Transcription factor footprinting analysis

#### Differential analysis of TF footprints

For the transcription factor (TF) footprinting analysis, we retrieved all Hsapiens TFs in the Jaspar 2022 database using the MotifDb R package (version 1.40.0) and calculated their footprint scores in each individual ATAC-seq library using the TOBIAS package^39^ (version 2.17). Briefly, to correct for bias in sequence preference of the Tn5 transposase, we used the ATACorrect function on the uniquely aligned non-mitochondria reads. Next, we used the ScoreBigWig function to calculate the footprint scores and the BINDetect function to assign TFs to the ATAC peaks. We manually validated the validity of the TF footprints for each TF and excluded TFs with non-canonical shape of aggregated footprints such as MAZ (Supplementary Figure 5c). The resulting bindetect_results.txt files from each ATAC-library stored the number of ATAC peaks bound by each TF. We merged the files into a single data frame and applied a linear mixed model for differential analysis. The model incorporated covariates significantly correlated with the top PCs: lme(Footprint scores ∼ Diagnosis + Age + PMI + BA9 + BA38 + BA39/40 + BA44/45 + FRiP, random = 1|Subject). The coefficient of the Diagnosis term was regarded as the difference in TF binding score between ASD and control samples. Transcription factors with a Diagnosis term coefficient > 0.0068 and a p-value < 0.1 were considered to have increased footprints, while those with a coefficient < -0.0055 and a p-value < 0.1 were considered to have decreased footprints.

#### Associating TF expression with chromatin accessibility

We derived the logFC values of TF gene expression from cerebral cortex from Gandal et al.^19^ and plotted it against the difference in TF binding scores between ASD and control samples. We focused our downstream analyses on the TFs with differential gene expression in the ASD or Dup15q brains (FDR < 0.05).

To investigate whether the ATAC peaks bound by our TFs of interest were more likely to exhibit differential chromatin accessibility, we employed a bootstrap approach. For each TF, we first obtained the union set of ATAC peaks bound by it in each library and quantified the count of ATAC peaks that showed differential chromatin accessibility. This union set of ATAC peaks were regarded as the TF-bound ATAC peaks in all the downstream analyses. Next, to assess the statistical significance of this count, we constructed a null distribution by randomly sampling 1000 times the number of TF-bound peaks from all the consensus ATAC peaks, and quantified the count of differential ATAC peaks in each sampled set. P-values were derived from the comparison of the actual count to this null distribution.

### Heatmap of chromatin accessible regions

We performed sample-wise scaling of the CQN-normalized logRPKM values of the ATAC peaks using the scale function in R. The heatmaps were generated using the Heatmap function in the ComplexHeatmap R package (version 2.14.0).

### Protein-protein interaction network analysis

To assess the enrichment of protein-protein interactions (PPIs) among the TFs that showed differential footprint and gene expression, we used both the STRINGdb R package (version 11.5) and the DAPPLE online tool (version 0.19, https://cloud.genepattern.org/gp/pages/index.jsf). The input list included all the 16 TFs whose gene expression in the whole cortex of the ASD brains changed in the same directions as their footprints: BACH1, BATF, BATF3, BHLHA15, BNC2, FOS, FOSL1, FOSL2, JDP2, JUNB, MEF2B, NFE2, NFIL3, TEF, ZNF211 and CTCF. For the STRINGdb analysis, we set network_type = “physical” and the resulting network are depicted in Supplementary Figure 5f. For the DAPPLE analysis, we set Permute Length = 50 and also obtained a PPI network (Fig. 3d).

### Assessing the overlap between the target genes of TF-bound ATAC peaks and the TF regulons

To assess the regulatory role of TFs on their proximal and distal target genes, we determined the statistical significance of the overlap between the target genes of the TF-bound ATAC peaks and the genes within the scRNA-seq regulon of that TF using Fisher’s Exact Test. The scRNA-seq regulons were retrieved from Wamsley et al.^14^, and the background gene list used for comparison was set to be the intersection of genes within all the scRNA-seq regulons and genes that were expressed in the bulk brain tissue as reported in Gandal et al.^19^.

### Identification of cell types with gene expression most affected by reduced CTCF binding in ASD

To identify the cell types in which gene expression is most affected by reduced CTCF binding, we first computed z-scores for each gene across the 36 cell clusters in single-nucleus RNA-seq (snRNA-seq) data from ASD and control brains^14^. We then calculated the average expression per cell cluster, performed z-score normalization across cell types for each gene, and subsetted to down-regulated genes^19^ exhibiting reduced distal chromatin accessibility and CTCF binding. The average z-scores of these genes were computed for each cell cluster, and statistical significance of the enrichment was assessed using Student’s t-test.

To determine whether cell types with the highest number of down-regulated genes in ASD were also among the most affected by reduced CTCF binding, we performed an enrichment analysis. The clusters with the highest number of down-regulated genes included ODC_1, EXT_6_L23, EXT_2_L56, EXT_9_L6, EXT_4_L56, INT_5_SST, ODC_2, EXT_3_L23. Five of these clusters rank among the top eight enriched for down-regulated genes with reduced distal chromatin accessibility and CTCF binding (Fig. 3g). The significance of this overlap was tested using Fisher’s exact test.

### Associating brain chromatin QTLs (caQTLs) with ATAC peaks

We obtained the 6200 brain caQTL-ATAC pairs from a previous bulk ATAC-seq study incorporating over 200 post-mortem brain samples^28^. We associated the brain caQTLs to our consensus ATAC peaks based on an overlap between our ATAC peaks and the ATAC peaks in Bryois et al.^28^, which yielded a final set of 3838 brain caQTL-ATAC pairs that were used for further analysis. Specifically, 179 caQTL-ATAC pairs were found to be associated with differential ATAC peaks associated with ASD.

### Associate rare non-coding SNVs with ATAC peaks

We obtained the list of *de novo* noncoding variants and their disease impact scores (DIS) in ASD probands and their unaffected siblings from a previous study on a cohort of 1790 ASD simplex families^50^. We extended the consensus ATAC peaks by 500bp and associated an ATAC peak to the noncoding variants if the extended peak region encompassed the variant. Using this approach, we assigned 9237 ATAC peaks to 9723 non-coding variants in ASD probands, and 9032 ATAC peaks to 9444 variants in unaffected siblings.

### Predicting the impact of SNVs on TF binding sites

We used the motifbreakR package^94^ (version 2.12.0) to predict the impact of SNVs on the binding motifs of transcription factors in the Jaspar 2022 Hsapiens database. We considered TF-disrupting SNVs as those with binding threshold of p-value < 1×10^-4^ and strong disruptive effects (alleleDiff < 0 and effect = “strong”). To assess the statistical significance of the difference in the proportion of TF-disrupting SNVs that were associated with ASD down-regulated ATAC peaks between ASD probands and unaffected siblings, we performed 1000 times of sampling. Each time we randomly selected 20 SNVs from the proband set and 20 SNVs from the sibling set, and counted the number of TF-disrupting SNVs in each set. We then compared the distributions of these two counts using the Wilcoxon Rank Sum test.

### CRISPR/Cas9-mediated deletion of regulatory regions and genetic variants

We selected three genomic loci containing ASD risk genes for validation, one associated with a caQTL and the other two associated with *de novo* non-coding single nucleotide variants (SNVs). For each genomic locus, we designed pairs of sgRNAs flanking a region of 1000-2000 bp in the ATAC peaks using the CRISPOR online tool (http://crispor.tefor.net/). In addition, we designed pairs of sgRNAs flanking the two SNVs, which locate within 500bp of the corresponding ATAC peaks. The sgRNAs on each side of the targeted regions were cloned into pL-CRISPR.EFS.tRFP (Addgene, 57818) and pL-CRISPR.EFS.GFP (Addgene, 57819) lenti-viral vectors respectively using the NEBridge Golden Gate Assembly protocol with BsmBI-v2 (NEB E1602). Virus was generated by co-transfection of 9 µg pL-CRISPR vectors with 900 ng pVSVg (Addgene, 8454) and 9 µg psPAX2 (Addgene, 12260) in HEK293 cells at 70-80% confluency.

Empty vectors without any sgRNA insertion were used as control. We combined the virus harvested from the cell media at 24 and 48 hours post-transfection and concentrated the virus using Lenti-X concentrator (Takara, 631232). Primary human neural progenitor cells (phNPCs) were seeded in 12-well plates and infected with a pair of viruses (5 µl/well of each concentrated virus) using protaminesulphate as helper (final concentration 8 µg/ml). 5 days after the infection, cells that were infected by both sgRNAs (RFP+/GFP+) were collected by FACS. Genomic DNA and RNA were extracted using the AllPrep DNA/RNA Micro kit (Qiagen, 80204), and cDNA were subsequently generated using the QuantiTect Reverse Transcription kit (Qiagen, 205311). Target gene expression level was measured by qRT-PCR using the iTaq Universal SYBR Green Supermix kit (Bio-Rad, 1725120) on QuantStudio 6 (ThermoFisher, 4485691) and the Design & Analysis Software (version 2.6.0, ThermoFisher). All gene expression was normalized to *GAPDH*. Each experiment was repeated 3-4 times, each time with 3-4 biological replicate wells of phNPCs and 3 technical replicate wells in qRT-PCR. The sgRNA and primer sequences for both genomic DNA and qRT-PCR were provided in Supplementary Table 10.

### Hi-C library preparation

To generate Hi-C libraries, we used the Arima Hi-C Kit (Arima Genomics, San Diego) according to the manufacturer’s instructions (Doc A160126 v01). To determine the input amount, we performed enzyme D treatment and DNA purification on 115k unsorted and 280k FANS-sorted nuclei, which yielded an estimate of 5-6 µg DNA per million nuclei. We calculated that 125k - 1M nuclei would satisfy the requirement of 750ng - 5µg DNA input. Around 500k nuclei from ASD and their matched control samples were used to generate bulk Hi-C libraries, while around 470k NeuNp and 880k NeuNn nuclei from Dup15q and their matched control samples were used to generate cell-type specific Hi-C libraries. The DNA concentration of each Hi-C library was quantified by the KAPA Library Quantification Kit (Ca. #KK4854) prior to library pooling and sequencing to an average depth of 865M reads per sample on the Illumina NovaSeq platform at 2×150 bp.

### Hi-C data processing and generation of interaction matrix

Paired-end sequencing reads were demultiplexed and processed using the HiC-Pro pipeline^95^ (version 3.1.0). Default settings were used to align reads to the human hg19 reference genome, remove duplicate reads, assign reads to restriction fragments produced by the cocktail of enzymes used in the Arima Hi-C Kit, filter for valid interaction pairs, and generate binned genome-wide interaction matrices at 10kb, 40kb, 100kb, and 250kb resolutions. Valid interaction pairs of the same Hi-C library were merged before generating the raw interaction matrices. For ASD and their matched control samples, the raw matrices were normalized using ICE (iterative correction and eigenvector decomposition) technique in the HiC-Pro pipeline. For Dup15q and their matched control samples, to correct for unwanted effects in the presence of CNVs, we applied CAIC and LOIC normalization^96^ (https://github.com/nservant/cancer-hic-norm) on the raw interaction matrices.

### Quality control of Hi-C libraries

We obtained the number of valid interactions, valid interactions after removal of duplicates, trans interactions, cis interactions, cis short -range interactions, and cis long-range interactions from the QC metrics compiled by the HiC-Pro pipeline. For each Hi-C library, we ensured that 1) over 35% of the valid interactions were classified as cis-long range interactions (loop distance > 20kb); 2) over 90% of the cells in the raw interaction matrix at 10kb resolution exceed 1000 contact counts. To assess the reproducibility between biological replicates of each cell type and diagnosis group, we calculated the Pearson’s correlation of the contact counts in the raw interaction matrix at 100kb resolution between each pair of samples within the group. All the within group correlations exceeded 0.82, demonstrating a high degree of reproducibility.

### Aggregation of Hi-C interaction maps

We first extracted the contact counts of intra-chromosomal interactions for each chromosome from the normalized Hi-C interaction matrix of each sample. Next, we grouped the samples by diagnosis and tissue or cell types and summed up the contact counts across samples within the same group. A 100kb resolution was used for comparing compartment assignments across cell types, and a 40kb resolution was used for all other occasions.

### A/B compartment analysis

Compartment scores were computed from the normalized Hi-C interaction matrix of each sample at 100kb and 250kb resolutions respectively using the HiTC R package^97^ (version 1.30.0, R version 3.6.1). Genomic bins with positive compartment scores were classified as the A compartments, while those with negative compartment scores as the B compartments.

To assign the similarity in compartment assignments across bulk, NeuNp and NeuNn samples, A/B compartments were called from the aggregated Hi-C interaction matrices at 100kb resolution of each group. Then we used the UpsetR package (version 1.4.0) to assess the overlap of genomic bins classified as A/B compartments between each group.

To assess the enrichment of cell-type specifically expressed genes within cell-type specific A compartments, we defined neuron-specific A compartments as the genomic bins at 100kb resolution that were classified as A compartment in all neurotypical NeuNp samples but as B compartment in all neurotypical NeuNn samples. Likewise, glia-specific A compartments were defined using comparable criteria.

### TAD analysis

We calculated the insulation scores and called TADs from the normalized Hi-C interaction matrix of each sample at 40kb resolution using the GENOVA R package^98^ (version 1.0.0, https://github.com/robinweide/GENOVA, R version 4.0.2) with parameters W = 16 and Min_strength = -1. We have tested a range of values for each parameter and selected the parameters with TAD calls that aligned best with the visual patterns observed in the Hi-C interaction maps. In addition, to compare the TAD structures between diagnosis and tissue or cell type groups, TADs were called from the aggregated Hi-C interaction matrices of each group (grouped by diagnosis and tissue or cell type). Then we used the UpsetR package (version 1.4.0) to assess the overlap between the TAD boundaries from each group.

### Differential insulation score analysis

For each tissue or cell type (bulk, NeuNp, NeuNn), we compiled TADs called from the respective disease group and their matched control group. Differential analysis was performed separately for each condition on insulation scores of 40kb bins around the TAD boundaries using linear mixed models, incorporating covariates that displayed significant correlation with the top PCs that explain at least 80% of the total variance. The model used for bulk tissue was: lme(Insulation score ∼ Diagnosis + Sex + Batch + Temporal lobe + Brain bank + Valid interaction% + Cis-long-range interaction%, random = ∼ 1|Subject). The model used for NeuNp nuclei was: lme(Insulation score ∼ Diagnosis + Sex + Valid interaction% + Duplicate% + Cis-long-range interaction%, random = ∼ 1|Subject). The model used for NeuNn nuclei was: lme(Insulation score ∼ Diagnosis + Age + BA7 + Duplicate% + Cis interaction%, random = ∼ 1|Subject). The coefficient of the Diagnosis term in the linear model was used as the logFC value, and those with FDR-corrected p-values of < 0.05 (idiopathic ASD) or 0.1 (Dup15q) were considered as significantly differential loops in each tissue or cell type.

### Statistical assessment of weakened TAD boundaries

For each tissue or cell type, we permuted the sample diagnosis labels (disease vs. control) 1000 times. Within each permutation, we conducted differential analysis on the insulation scores using the same aforementioned linear mixed models and quantified the difference in the number of weakened versus strengthened TAD boundaries, while adhering to the specified FDR thresholds. Subsequently, we determined the fraction of permutations in which the quantified difference surpassed our observed value and use this fraction as the p-value of our statistical analysis.

### Chromatin loop analysis

#### Chromatin loop calling

Chromatin loops were called for each sample using the FitHiC2 python package^99^ (version 2.0.7, https://github.com/ay-lab/fithic, python version 3.6.8) with the raw Hi-C interaction matrix and biases produced by HiC-Pro at 10kb resolution as input, and a q-value cutoff of < 0.01 was applied.

#### Identification of consensus loops and comparison between tissue or cell types

Consensus chromatin loops for each tissue or cell type were defined as those that were called in at least 8 out of the 36 bulk cortical samples, 9 out of the 15 NeuNp samples, and 7 out of the 15 NeuNn samples, respectively. These sample thresholds were chosen based on visual evaluation of randomly sampled loci. Specifically, we identified 34,657 consensus chromatin loops in bulk cortical samples, 38,142 in NeuNp samples, and 46,180 in NeuNn samples.

The consensus loop sets were used to compare chromatin loop interactions across different tissue or cell types. To ensure robustness, we filtered out loops that anchored at segmental duplicate regions, using the SeqDup_build37.txt file downloaded from the UCSC genome browser. Moreover, we restricted our analysis to loops with distances ranging from 20kb to 5Mb. Then we used the UpsetR package (version 1.4.0) to assess the overlap between the three loop sets.

#### Classification of loop types

To classify the loop interaction types, we defined promoters as the region between 2kb upstream and 100bp downstream of the TSSs of all the known genes in hg19 (TxDb.Hsapiens.UCSC.hg19.knownGene), and enhancers as the ATAC consensus peaks that did not overlap with the promoters. Among the consensus loops, those that had both loop ends intersect the promoters were classified as promoter-promoter interactions (P-P), those that had both loop ends intersect the enhancers were classified as enhancer-enhancer interactions (E-E), and those that had one loop end intersect the promoters and the other end intersect the enhancers were classified as promoter-enhancer interactions (P-E). The other loops were classified as None. Note that one loop can be classified into multiple categories, as the 10kb loop end may overlap both a promoter and an enhancer. Therefore, the sum of these four categories could exceed 100%.

#### Cell-type specific promoter loops

Neuron-specific loops were defined as the consensus loops in NeuNp samples that were never called as a chromatin loop in any NeuNn sample. Likewise, glia-specific loops were defined as the consensus loops in NeuNn samples that were never called as a chromatin loop in any NeuNp sample. Then we designated cell-type specific promoter loops as those that had at least one loop end encompassing a gene TSS.

#### Differential loop analysis

For differential loop analysis, we first calculated the sample-wise Count Per Million (CPM) value for each consensus loop by dividing the number of contact counts of that loop by the total number of contact counts of all the consensus loops in million, and then took the logarithm of the CPM (logCPM) with a prior count of 0.5. This logCPM value was used to build linear mixed models, incorporating covariates that displayed significant correlation with the top PCs. The model used for bulk tissue was: lme(Loop logCPM ∼ Diagnosis + Sex + Batch + BrainBank Harvard-ATP + BA 39/40 + BA 44/45 + Valid interaction% + Trans interaction%, random = ∼ 1|Subject). The model used for NeuNp nuclei was: lme(Loop logCPM ∼ Diagnosis + Sex + BA 4/6 + Valid interaction% + Trans interaction%, random = ∼ 1|Subject). The model used for NeuNn nuclei was: lme(Loop logCPM ∼ Diagnosis + Age + PMI + Duplicate% + Cis-long-range interaction% + Cis-short-range interaction% + cis-short-range interaction%, random = ∼ 1|Subject). The coefficient of the Diagnosis term in the linear model was used as the logFC value, and those with FDR-corrected p-values of < 0.05 were considered as significantly differential loops in each tissue or cell type.

### FIRE analysis

#### FIRE calling

FIREs were called for each sample using the FIREcaller R package^100^ (version 1.40, https://github.com/yycunc/FIREcaller, R version 4.0.2) on the raw Hi-C interaction matrix at 40kb resolution. The mappability file customized for Arima-HiC was downloaded from https://github.com/HuMingLab/MAPS/tree/master/Arima_Genomics/genomic_features. We defined consensus FIREs in each cell type as the 40kb genomic bins that were called as FIREs (the 6^th^ column indicator =1 in the FIRE output file) in at least 20% of the samples.

#### Differential FIRE analysis

We restricted our analysis to the union of consensus FIREs in bulk tissue, NeuNp and NeuNn nuclei, which summed up to 7,582 bins, each at 40kb in length. Differential FIRE analysis was performed using the nlme R package (version 3.1.160) on each tissue or cell type separately by fitting a linear mixed model for the FIRE scores (the 4^th^ column named norm_cis in the FIRE output file of each sample), incorporating covariates that displayed significant correlation with the top PCs. The model used for bulk tissue was: lme(FIRE scores ∼ Diagnosis + Age + Sex + PMI + Batch + BrainBank NICHD-BTB + Temporal cortex + Valid interaction% + Cis-long-range interaction%, random = ∼ 1|Subject). The model used for NeuNp nuclei was: lme(FIRE scores ∼ Diagnosis + Frontal cortex + Valid interaction% + Trans interaction%, random = ∼ 1|Subject). The model used for NeuNn nuclei was: lme(FIRE scores ∼ Diagnosis + Age + Sex + Valid interaction% + Trans interaction% + Cis-short-range interaction%, random = ∼ 1|Subject). The coefficient of the Diagnosis term was used as the magnitude of change in FIREs, and those with FDR-corrected p-values of < 0.05 were considered as significantly differential FIREs.

#### Associate the changes in FIREs between diagnosis and with other modalities

To assess the relationship of FIRE changes between idiopathic ASD and Dup15q syndrome, and across tissue and cell types, we calculated the Pearson’s correlation between the magnitude of change in FIREs. We calculated the correlations using either 1) all 7,582 FIREs or 2) limited to FIREs outside of the Dup15q region (chr15:20,000,000-33,000,000). Both were significant, and the correlation coefficients and p-values of the latter analysis are shown in the figures. To investigate the relationship between alterations in FIRE and changes in ATAC signal in bulk ASD samples, we used the 7,164 (out of 7,582) FIREs that overlapped with 25,697 consensus ATAC peaks. Pearson’s correlation was calculated between the magnitude of change in FIREs and the ATAC logFC values. In addition, to investigate the relationship between changes in FIRE and chromatin loop interaction in bulk ASD samples, we conducted the analysis on 12,720 consensus chromatin loops in bulk cortical tissue that had at least one loop end within the 7,582 FIREs. Specifically, 3249 FIREs were involved. Pearson’s correlation was calculated between the magnitude of change in FIREs and the loop interaction logFC values.

### Enrichment of cell-type specifically expressed genes

We downloaded the gene expression profiles of neurons, astrocytes, oligodendrocytes, endothelial cells, and microglia from Zhang et al^101^ and computed the average expression in FPKM for each gene in each cell type. To identify the cell type marker genes, or cell type specifically expressed genes, we calculated the cell-type specificity index of each gene using the pSI R package^102^ (version 1.1, http://genetics.wustl.edu/jdlab/psi_package/). To assess the enrichment of these cell type marker genes in our gene lists of interest, we calculated the odds ratio (OR) and the corresponding p-values using Fisher’s Exact Test. To account for multiple testing, we applied the False Discovery Rate (FDR) approach to adjust the p-values across all the gene lists and cell types. We used a cutoff of OR > 1 and FDR < 0.1 to determine statistically significant enrichment.

### Distance between cell-type specifically expressed genes

We used the EnsDb.Hsapiens.v75 R package (version 2.99.0) to retrieve the most upstream start coordinate of transcripts for each cell-type specifically expressed gene. We excluded transcripts with biotypes “retained_intron” or “non_stop_decay”, as well as pseudogene transcripts, from the analysis. Next, we calculated the distance between the transcript start coordinates of each pair of adjacent genes on the same chromosome for each of the five cell types (endothelia, neuron, microglia, astrocyte, oligodendrocyte).

### Subtraction of Hi-C interaction matrices

To investigate the chromosome structure of the duplicated region in the Dup15q samples, we subtracted the aggregated Hi-C matrix of the cell-type-matched control samples from each of the individual Dup15q matrices. We restricted the analysis to intra-chromosomal interactions within chromosome 15 at 40kb resolution. Prior to performing the subtraction, we ensured that the total contact counts of both the control aggregated matrix and the individual Dup15q matrix were adjusted to 1 billion using the load_contacts function in the GENOVA R package. Then we performed the subtraction operation on the two lists of contacts.

### Differential analysis on DNA methylation

We obtained the wateRmelon-normalized methylation beta values for 417,460 probes across the genome in 74 ASD and 42 control samples from 42 ASD and 27 control brains from Wong et al^66^. Our analysis focused on CpG islands, and for each CpG island, we calculated the average of the normalized methylation beta values across all relevant probes (excluding the probes at CpG shores or shelves). We fit linear mixed models for the averaged normalized methylation beta values, incorporating all covariates that displayed significant correlation with the top PCs (FDR < 0.05). Colinearity among the covariates was evaluated using the olsrr package (version 0.5.3) and yielded a variance inflation factor (VIF) of < 2. We treated technical replicates as a random effect in the linear mixed model: lme(Average of normalized beta values ∼ Diagnosis + CET + Age + BA9 + Sex + Batch + DilPlate2 + Call.rate + Brainbank_NICHD, random = ∼ 1|Subject), where CET score represents the proportion of neuronal vs glial cells^21,103^. CpG islands with an FDR-corrected p-value of < 0.05 between the ASD and control samples as differentially methylated regions (DMRs).

### Enrichment of hypermethylated CpG islands at weakened TAD boundaries

We assessed the enrichment of hypermethylated CpG islands at the 754 weakened TAD boundary bins by comparing the percentage of hypermethylated CpG islands within these bins to that observed in randomly sampled TAD boundary bins. To establish a null distribution, we employed a bootstrap approach. In each iteration, 754 bins were randomly selected from a total of 11167 bins (bin size: 40kb) surrounding the TAD boundaries, and the percentage of hypermethylated CpG islands within these bins was calculated. This process was repeated 500 times. In only 2% of iterations did the percentage from the null distribution exceed the observed percentage.

### Associate brain methylation QTLs (mQTLs) with ASD disease risk

We obtained a dataset comprising 16809 fetal brain mQTLs from a DNA methylation study on 166 human fetal brain samples^67^, using the same HumanMethylation450 beadarray platform as the DNA methylation data we used from Wong et al.^66^. Notably, the study demonstrated a strong correlation between mQTL effect sizes observed in adult brain regions and those observed in fetal brains. Of the 16809 fetal mQTLs, 8057 were found to overlap the ASD GWAS summary statistics^72^. Among these overlapping mQTLs, 3344 were associated with CpG islands and 46 locate within DMRs at weakened TAD boundaries.

### Individual contributions of chromatin accessibility change and weakened TAD boundaries

We used an FDR threshold of 0.05 to designate differentially expressed genes (DEGs) in the cerebral cortex of idiopathic ASD samples, using the whole cortex data from Gandal et al^19^. To identify DEGs associated with differential ATAC peaks (FDR < 0.1), we required that the gene expression change should align with the direction of ATAC peak changes. Additionally, the gene should be predicted as a target gene of the ATAC peak following the approach detailed earlier. To identify DEGs associated with weakened TAD boundaries in idiopathic ASD, we included all DEGs that overlap with the 40kb bins exhibiting increased insulation scores and meeting an FDR threshold of 0.1.

## 2. Supplementary Figures

**Supplementary Figure 1 | Fragment size distribution of ATAC-seq libraries a,** Quality assessment of the ATAC-seq libraries based on their fragment size distributions. Only the “Good” library quality samples were used for downstream analysis.

**Supplementary Figure 2 | Build linear model for differential ATAC analysis a,** Correlation of the top PCs of the cqn-normalized ATAC peak read count matrix with the biological and technical covariates. **b,** Correlation of the biological and technical covariates with themselves.

**Supplementary Figure 3 | Differential chromatin accessibility and target gene expression a,** Distribution of p-values obtained from the linear mixed model, which was used to assess differential chromatin accessibility between ASD and neurotypical brains. **b,** Venn diagram showing significant overlap between the number of base pairs in differential ATAC peaks in ASD brains and those in differential H3K27ac peaks from a previous study (Ramaswami et al, 2020 Nat Commun). **c,** Violin plot showing increased correlation of ATAC signal between pairs of ATAC peaks linked by consensus Hi-C chromatin loops in bulk tissue. **d**, Scatterplot showing significant correlation between the log fold changes in promoter ATAC signal and proximal gene expression **e**, Scatterplot showing correlation between the log fold changes in distal ATAC signal and their target gene expression.

**Supplementary Figure 4 | Distribution of DARs in different developmental stages and cell types a,** Number of ATAC peaks that are open chromatin regions in adult or fetal brains. On the left are known ATAC peaks compiled from the literature (Bryois et al., 2018, Nat Commun; Torre-Ubieta et al., 2018, Cell; Markenscoff-Papadimitriou et al., 2020, Cell). On the right is a breakdown of the number of DARs in each category. Open chromatin regions that overlap between the two developmental stages are labeled as constitutive. **b,** Cell type specificity of the DARs. On the left are numbers of ATAC peaks that are reproducible ATAC peaks in each single cell cluster or cell type. On the right is a breakdown of the number of DARs overlapping these reproducible ATAC peaks. **c,** Scatterplots showing correlation of the chromatin accessibility changes between DARs in bulk tissue and in each single cell clusters. Only the ATAC peaks that are differential (FDR < 0.05) in both the bulk tissue and the corresponding single cell cluster are shown in each mini-plot.

**Supplementary Figure 5 | Transcription factor footprint, gene expression and physical interaction network a-c,** Aggregated footprint of CTCF (a), BATF (b), or MAZ (c) across all the samples. **d,** Scatterplot showing differential gene expression of the TFs that show differential binding scores between ASD and CTL. The TFs with significantly dysregulated gene expression in brains of individuals with Dup15q syndrome are highlighted in red (FDR < 0.05). **e,** Histograms showing the distributions of the expected numbers of dysregulated ATAC peaks bound by the TFs of interest. The observed numbers are shown by the red bars. **f,** STRING protein-protein interaction network showing physical interactions among TFs whose gene expression and binding scores increase in ASD brains.

**Supplementary Figure 6 | Transcription factors driving dysregulation of chromatin accessibility and transcriptomic changes a**, Scatterplot showing significant correlation between the log fold changes in BACH1-bound promoter ATAC intensity and their proximal gene expression **b**, Scatterplot showing no significant correlation between the log fold changes in BACH1-bound distal ATAC intensity and their target gene expression. **c**, Scatterplot showing significant correlation between the log fold changes in CTCF-bound promoter ATAC intensity and their proximal gene expression **d,** Venn diagram showing significant overlap between the target genes of BACH1-bound promoter ATAC peaks and genes in the BACH1 regulon in a scRNA-seq dataset (Wamsley et al.). **e,** Venn diagram showing significant overlap between the target genes of CTCF-bound distal but not promoter ATAC peaks and genes in the CTCF regulon in a scRNA-seq dataset (Wamsley et al.). **f**, Venn diagram showing significant overlap between the ASD up-regulated genes with promoter ATAC bound by BATF, FOSL1, BACH1, or NFE2 and the M9 astrocyte module in Parikshak et al. 2016 (Parikshak et al. 2016).

**Supplementary Figure 7 | Effect of brain caQTLs of ASD non-differential ATAC peaks on disease risk c,** Scatterplot showing no correlation between the effect of brain caQTLs on non-differential distal ATAC peaks and their effect on ASD disease risk.

**Supplementary Figure 8 | Validation of SNP and SNV effect on ASD risk genes using CRISPR/Cas9-mediated deletions a,** Schematic representation of locations of the targeted ATAC peak, SNP, and three CRISPR/Cas9-mediated genomic deletions at the locus shown in Fig. 4b. **b,** qRT-PCR results showing expression of the three genes *TRMT1, NFIX* and *NACC1* following CRISPR/Cas9-mediated deletion, in comparison to mock treatment with empty vectors, using *GAPDH* as the reference gene. *** denotes Dunnett-corrected p-value < 0.001, while “ns” denotes Dunnett-corrected p-value > 0.05. **c,** The SNV chr22:40576084:A:G identified in ASD proband (example in Fig. 4f) is predicted to disrupt binding of transcription factors SOX4 and SOX10. **d,** The SNV chr19:4033628:G:A identified in ASD proband (example in Fig. 4g) is predicted to disrupt binding of transcription factor ZNF135. **e,** Schematic representation of locations of the targeted ATAC peak, SNV, and the three CRISPR/Cas9-mediated genomic deletions at the locus shown in Fig. 4g. **f,** qRT-PCR results showing expression of the three predicted target gene *PIAS4* and two other genes *ZBTB7A* and *MAP2K2* following CRISPR/Cas9-mediated deletion, in comparison to mock treatment with empty vectors, using *GAPDH* as the reference gene. ** denotes Dunnett-corrected p-value < 0.01, **** denotes Dunnett-corrected p-value < 0.0001, while “ns” denotes Dunnett-corrected p-value > 0.05.

**Supplementary Figure 9 | Comparison of 3D genomic architecture in bulk and NeuN-sorted nuclei of post-mortem cortical samples a,** Contour and dot plot from FANS-sorting analysis showing gated NeuN-positive (neuronal) and NeuN-negative (glial) populations. The numbers and percentages of each population are summarized in the table. **b,** Scatterplot showing the number of NeuNp and NeuNn nuclei collected from FANS-sorting of postmortem brain cortical samples with different weights. **c,** Violin showing the distribution of the Pearson’s correlation of raw Hi-C matrices at 100 kb resolution, demonstrating a high degree of reproducibility (correlation coefficients all > 0.82) between biological replicates from different individuals within each cell type and diagnosis group. **d,** Enrichment analysis showing that genes within NeuNp and NeuNn-specific A compartments are significantly enriched for their corresponding cell-type specifically expressed genes. Enrichments with FDR < 0.1 are labeled with odds ratio followed by FDR values in the parentheses. **e,** Bar graph showing the percentage of consensus chromatin loops in each cell type between pairs of gene promoters (P-P), enhancers (E-E), enhancer-promoters (E-P) or other regions. Because the chromatin loops were identified at 10kb resolution, the loop ends may encompass multiple gene promoters and enhancers, and therefore the total percentage may exceed 100%.

**Supplementary Figure 10 | 3D genomic architecture at the Dup15q locus a,** Genomic tracks showing compartment scores and TAD structures at chr15:21,500,000-33,000,000 in samples with different diagnosis groups and cell types.

**Supplementary Figure 11 | Shared and distinct 3D genomic architecture across bulk and NeuN-sorted nuclei of post-mortem cortical samples a,** Upset plot showing the overlap of A compartment assignments between bulk, NeuNp and NeuNn nuclei. **b,** Upset plot showing the overlap of TAD boundaries between samples with different diagnosis groups and cell types. **c,** Cell-type specific Hi-C maps and bulk ATAC-seq track at the *GRIA1* locus showing NeuNp-specific TAD boundaries and promoter loops, in comparison to the NeuNn structures. TAD boundaries are marked by grey triangles, and location of the NeuNp-specific promoter loop is marked by the red circles and an arrow. **d,** Pie charts showing the proportion of consensus Hi-C loops unique to each cell type and shared between different cell types.

**Supplementary Figure 12 | Global decrease of TAD boundary strength in ASD and Dup15q post-mortem cortical samples a, c, e,** MA plot showing differential insulation score at TAD boundaries in ASD bulk (a), Dup15q neurons (c) or Dup15q glia (e) with FDR < 0.1. **b, d, f,** Histograms showing the distribution of the expected difference in the number of weakened and strengthened TAD boundaries in ASD bulk (b), Dup15q neurons (d) or Dup15q glia (e) using a permutation strategy. The observed numbers are shown by the red bar.

**Supplementary Figure 13 | Changes in chromatin loops and FIREs in idiopathic ASD and Dup15q syndrome a,** MA-plots showing disease differential promoter-enhancer loops (p < 0.05) in bulk or NeuN-sorted nuclei from postmortem brains of individuals with idiopathic ASD or Dup15q syndrome. **b,c,** Histograms showing the distributions of p-values obtained from the linear mixed models, which were used to assess differential promoter loops (b) and FIREs (c) between the disease and control samples in bulk or NeuN-sorted nuclei.

**Supplementary Figure 14 | Coordinated changes in 3D genomic architecture between idiopathic ASD and Dup15q syndrome and with differential chromatin accessibility a,** Scatterplot showing significant correlation of changes in the insulation scores between idiopathic ASD bulk tissue and Dup15q neurons (left) or glia (right). **b,c,** Scatterplot showing significant correlation between changes in FIRE scores and changes in promoter loops (b) or ATAC signal (c). **d,** Scatterplot showing significant correlation of changes in FIRE scores between idiopathic ASD bulk tissue and Dup15q neurons (left) or glia (right). The correlation coefficients and p-values were calculated using the FIREs outside of the Dup15q region (chr15: 20,000,000-33,000,000).

**Supplementary Figure 15 | Enrichment of ASD-associated differentially expressed genes (DEGs) within 15 Mb of TAD boundaries** Odds ratios and p-values of the enrichment of ASD-associated DEGs within 15 Mb of weakened (top row) or strengthened (bottom row) TAD boundaries in idiopathic ASD postmortem brains.

## 3. Supplementary Tables

**Supplementary Table 1 - Demographic features and biological covariates of idiopathic ASD, Dup15q syndrome, and control subjects for ATAC-seq and Hi-C.**

**Supplementary Table 2 - Significantly correlated distal-promoter ATAC peak pairs.** The “promoter_ATAC” and “distal_ATAC” columns contain the locations of the promoter and distal ATAC peaks. The “loop” column indicates the location of the chromatin loop that connects each distal-proximal ATAC peak pair. ATAC peak pairs within 30kb of distance is marked as “Within30kb” in this column. The “cor” column contains the sample-wise Pearson’s correlation of the cqn-normalized logRPKM values of each ATAC peak pair. The “FDR” column contains the FDR-corrected p-values of the Pearson’s correlations.

**Supplementary Table 3 – Distal ATAC-gene connections identified using the ABC model.**

**Supplementary Table 4 – Transcription factors enriched in DARs or exhibit differential footprinting.**

**Supplementary Table 5 - Brain chromatin QTLs associated with ASD-dysregulated ATAC peaks.** The “Effect_size_on_ATAC” column contains the slope of each effect allele on ATAC peak signal from Bryois et al.^28^. The “ASD_GWAS_Zscore” column contains the Z-score of each effect allele in the summary statistics of the ASD GWAS study^72^. The “ATACpeak” column indicates the locations of the ATAC peaks associated with each SNP. The “ATAC_logFC” and “ATAC_FDR” columns contain the log fold change and FDR-corrected p-values of each ATAC peak in the bulk ASD brain samples compared to the neurotypical CTL samples. Only the differential ATAC peaks with FDR < 0.05 are listed in this table. The “ATAC_target_gene_id” and “ATAC_target_gene_name” columns contain the Ensembl gene IDs and gene names of the target genes of the ATAC peaks. The “ATAC_type” column indicates whether each ATAC peak locates at the gene promoter region, which is defined as the region between 2kb upstream and 100bp downstream of the gene TSSs, or distal region of the corresponding target gene.

**Supplementary Table 6 – Relationship between the CRISPR-targeted region and potential target genes**

**Supplementary Table 7 - QC metrics of Hi-C libraries.**

**Supplementary Table 8 – Differentially methylated regions in ASD brains**

**Supplementary Table 9 – Contribution of promoter and distal chromatin accessibility changes and weakened TAD boundaries to differentially expressed genes in ASD brains.** In the four DAR columns, an association of DEG with is indicated by “Y”. In the weakened TAD boundaries column, a DEG is labeled “Y” if it is located within 40kb of the weakened TAD boundaries.

**Supplementary Table 10 – sgRNA and primer sequences for genomic DNA PCR and qRT-PCR used in the CRISPR validation experiments.**

## Notes

### Competing Interest Statement

The authors have declared no competing interest.

### Author Declarations

IRB of University of California Los Angeles gave ethical approval for this work.

