## Supplementary Figures for "Alterations in chromatin accessibility and conformation elucidate genetic mechanisms in ASD"

### Supplementary Figure 1

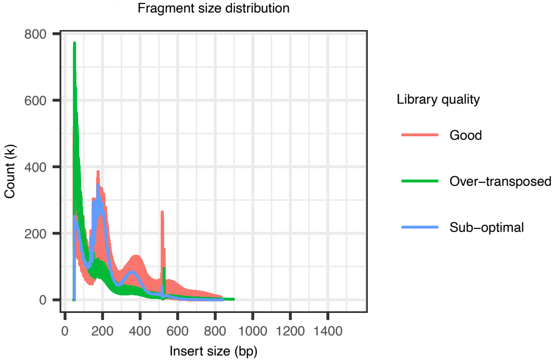

Supplementary Figure 2

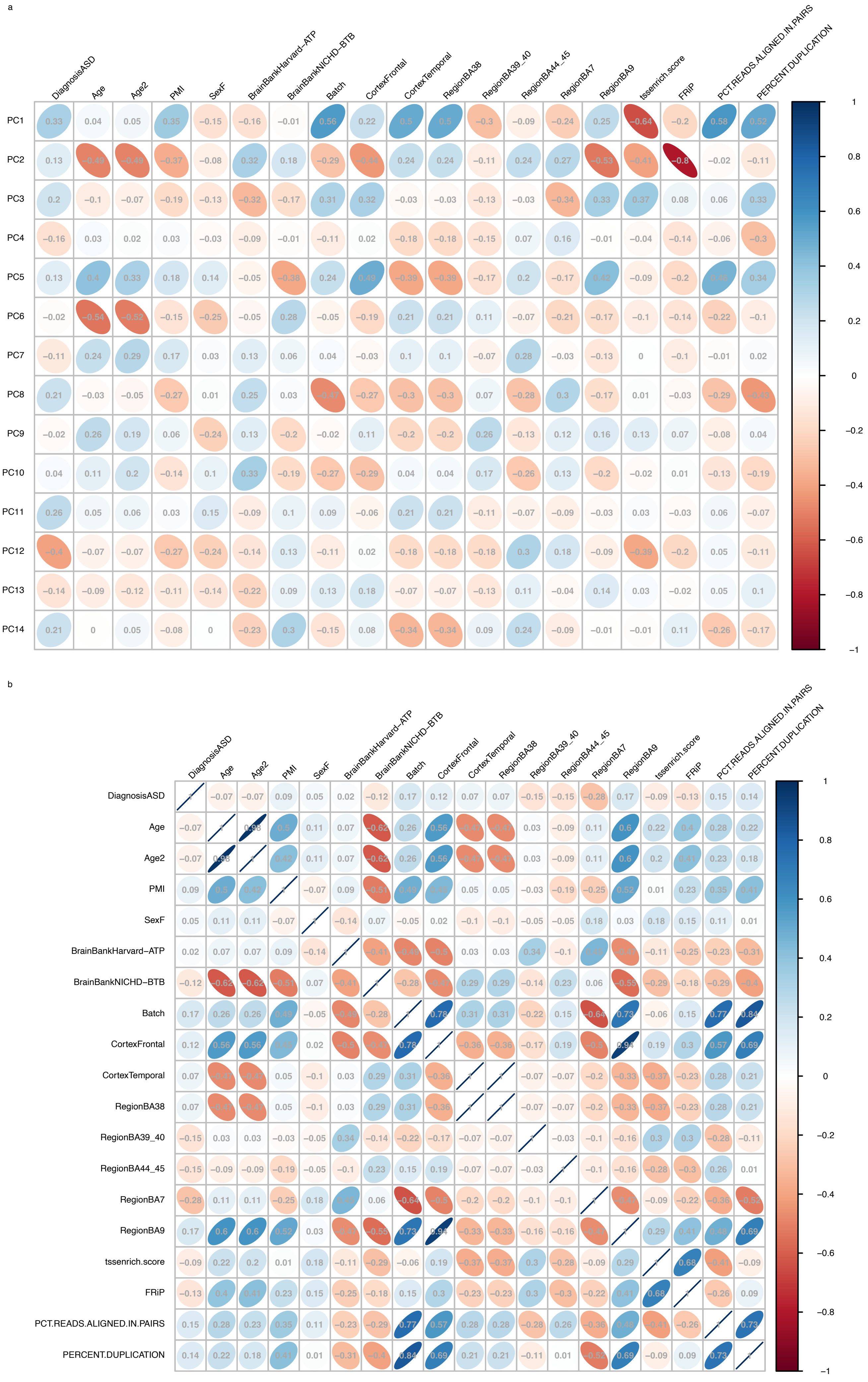

Supplementary Figure 3

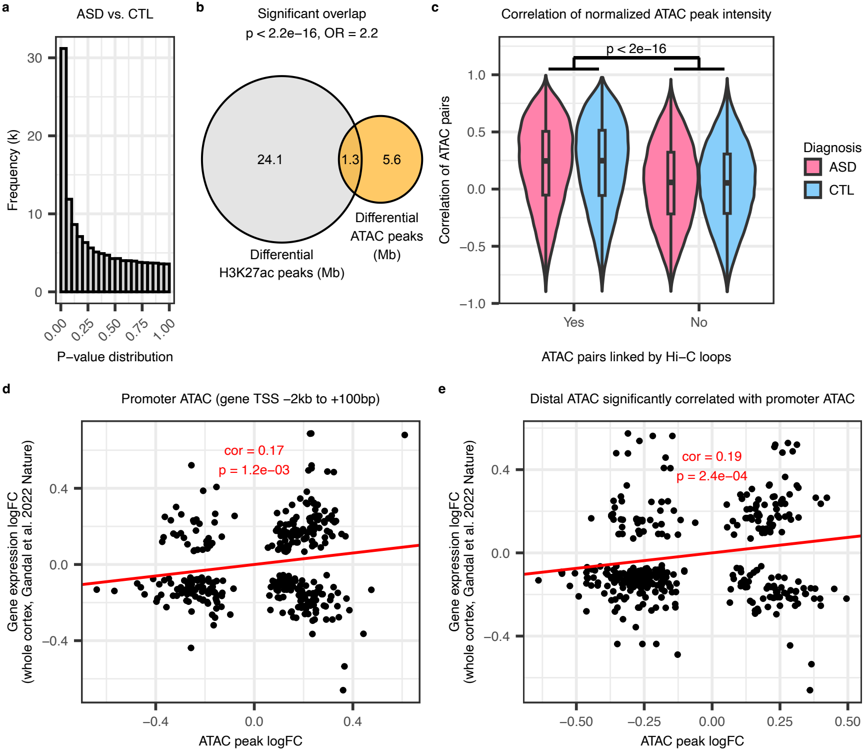

Supplementary Figure 4

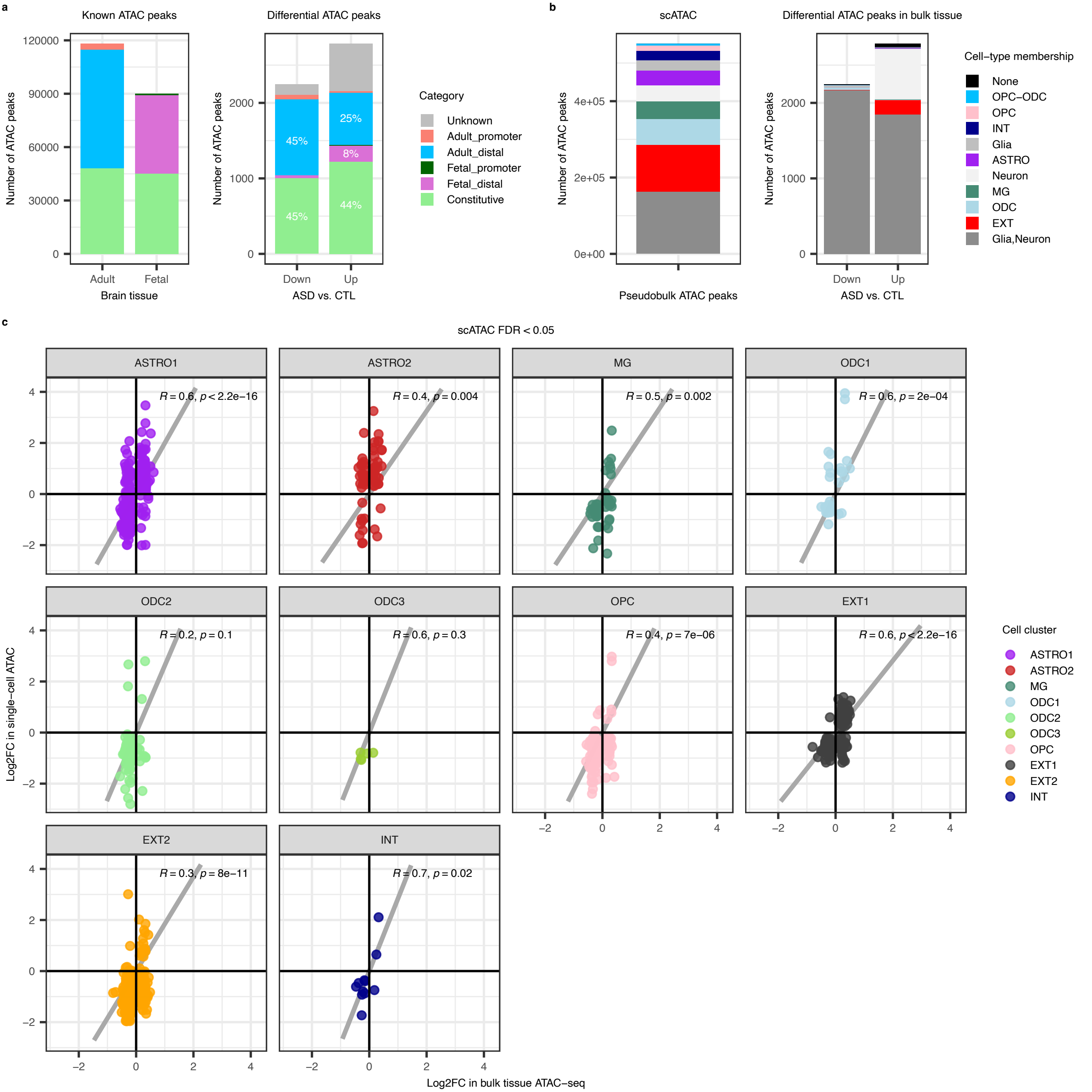

Supplementary Figure 5

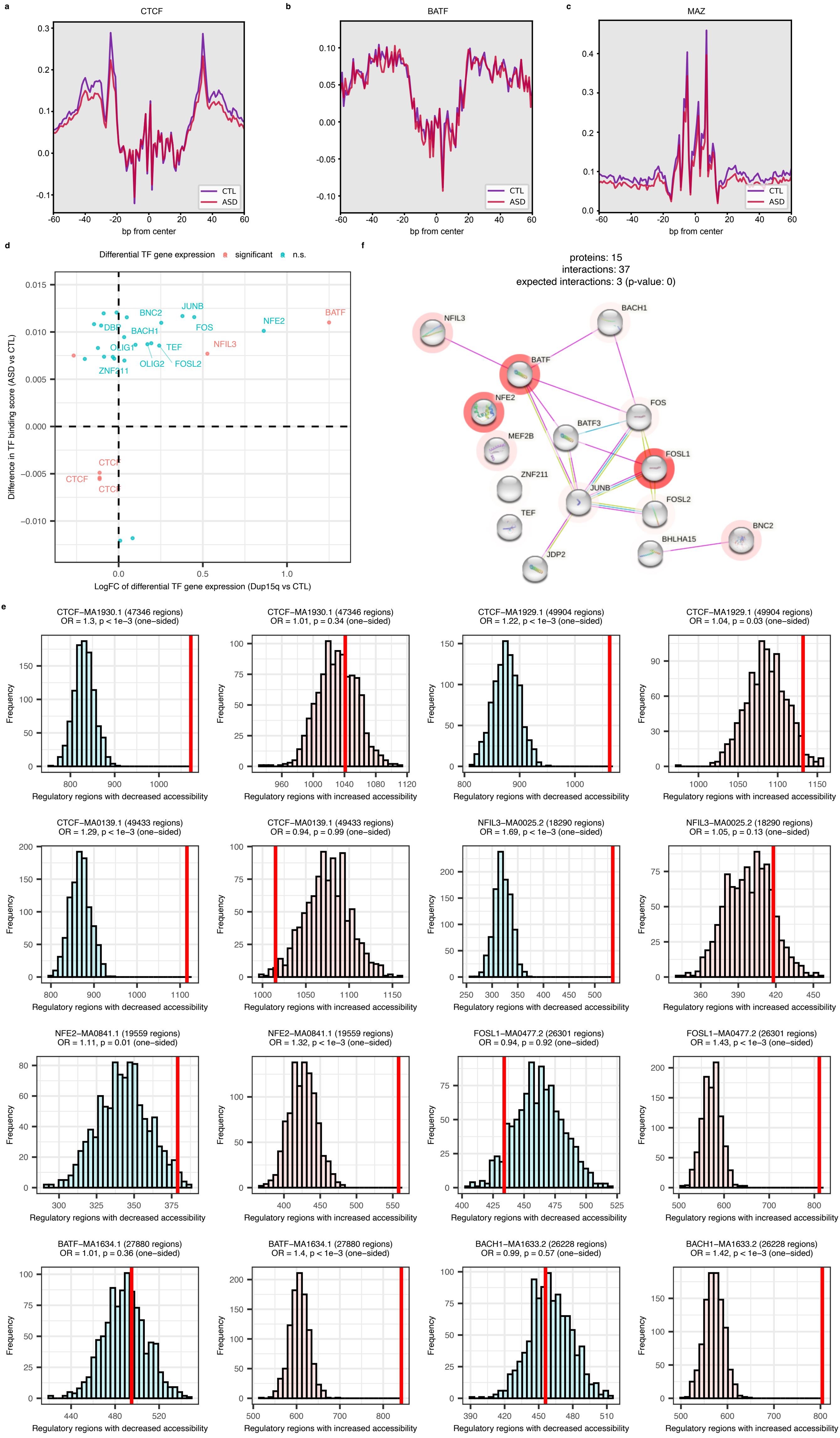

Supplementary Figure 6

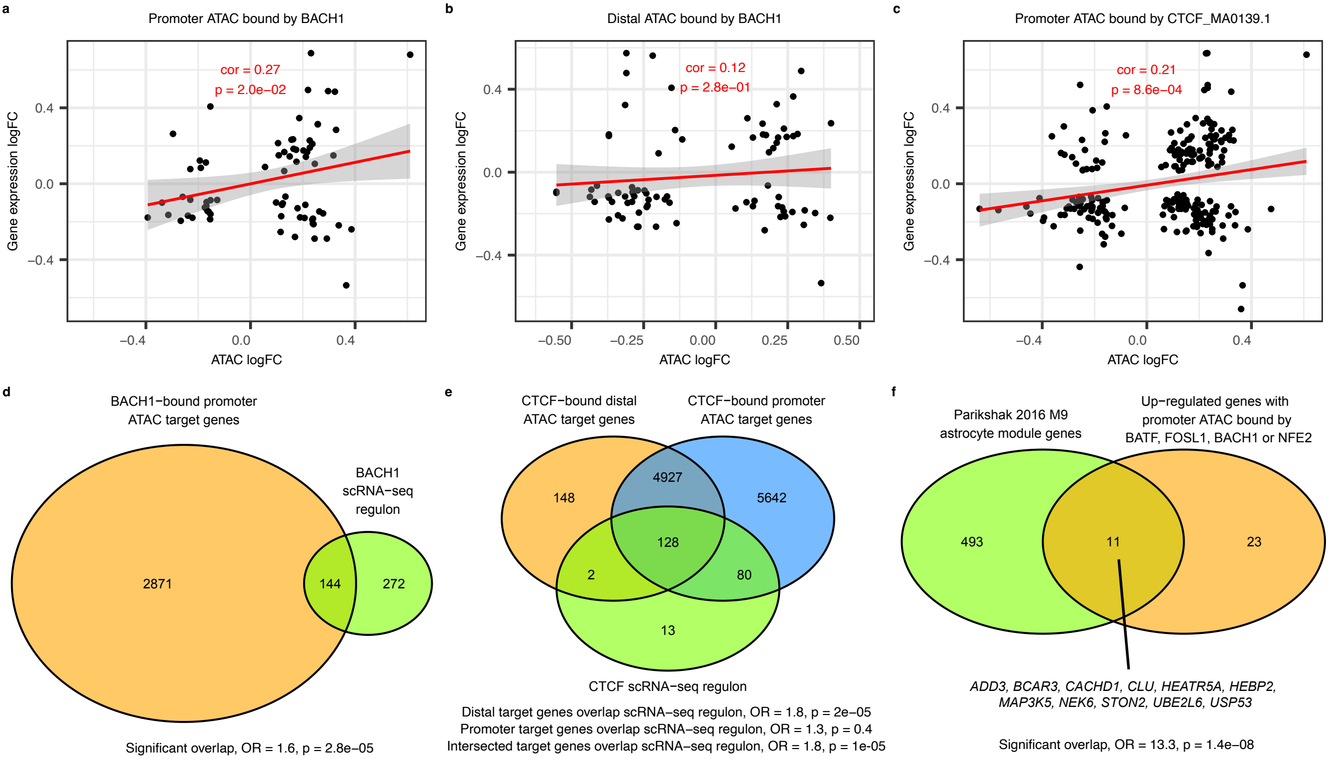

Supplementary Figure 7

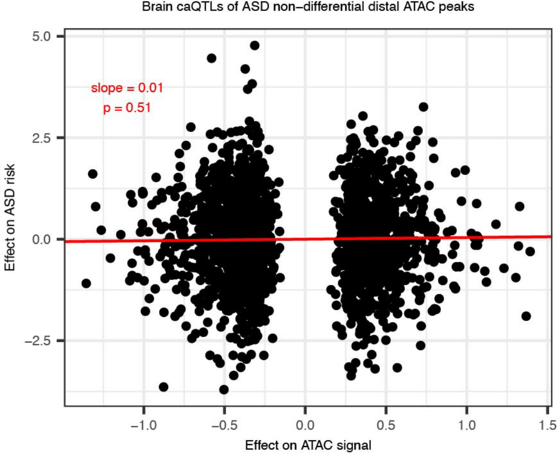

Supplementary Figure 8

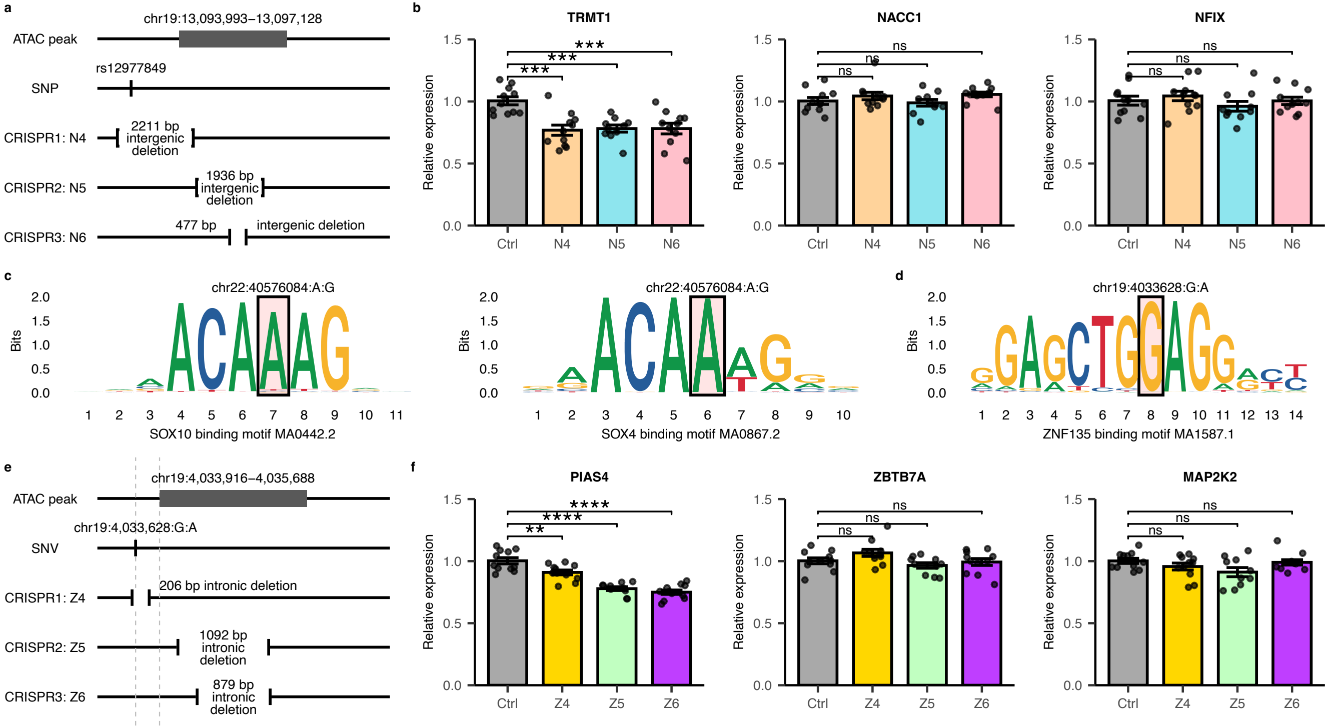

Supplementary Figure 9

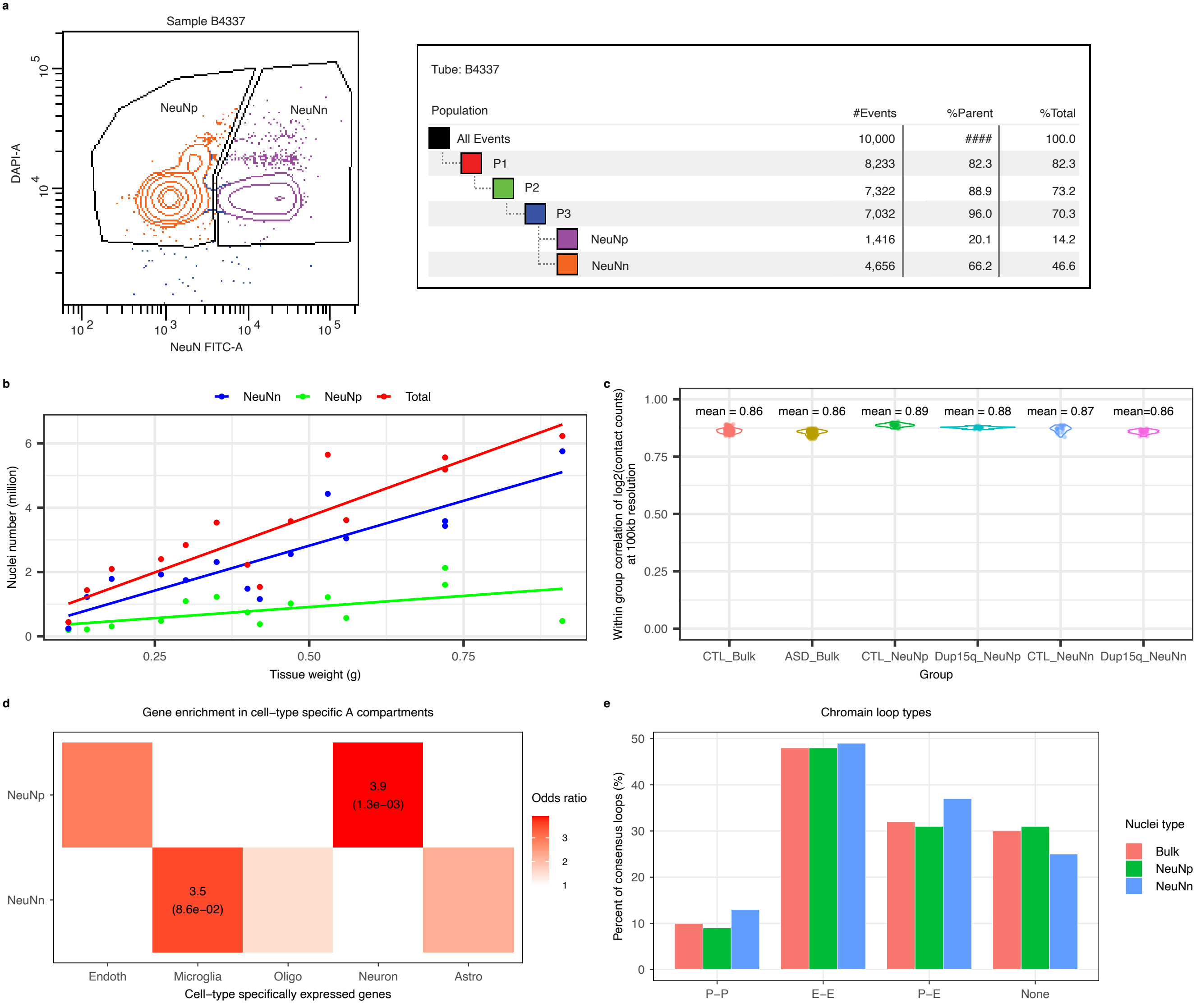

Supplementary Figure 10

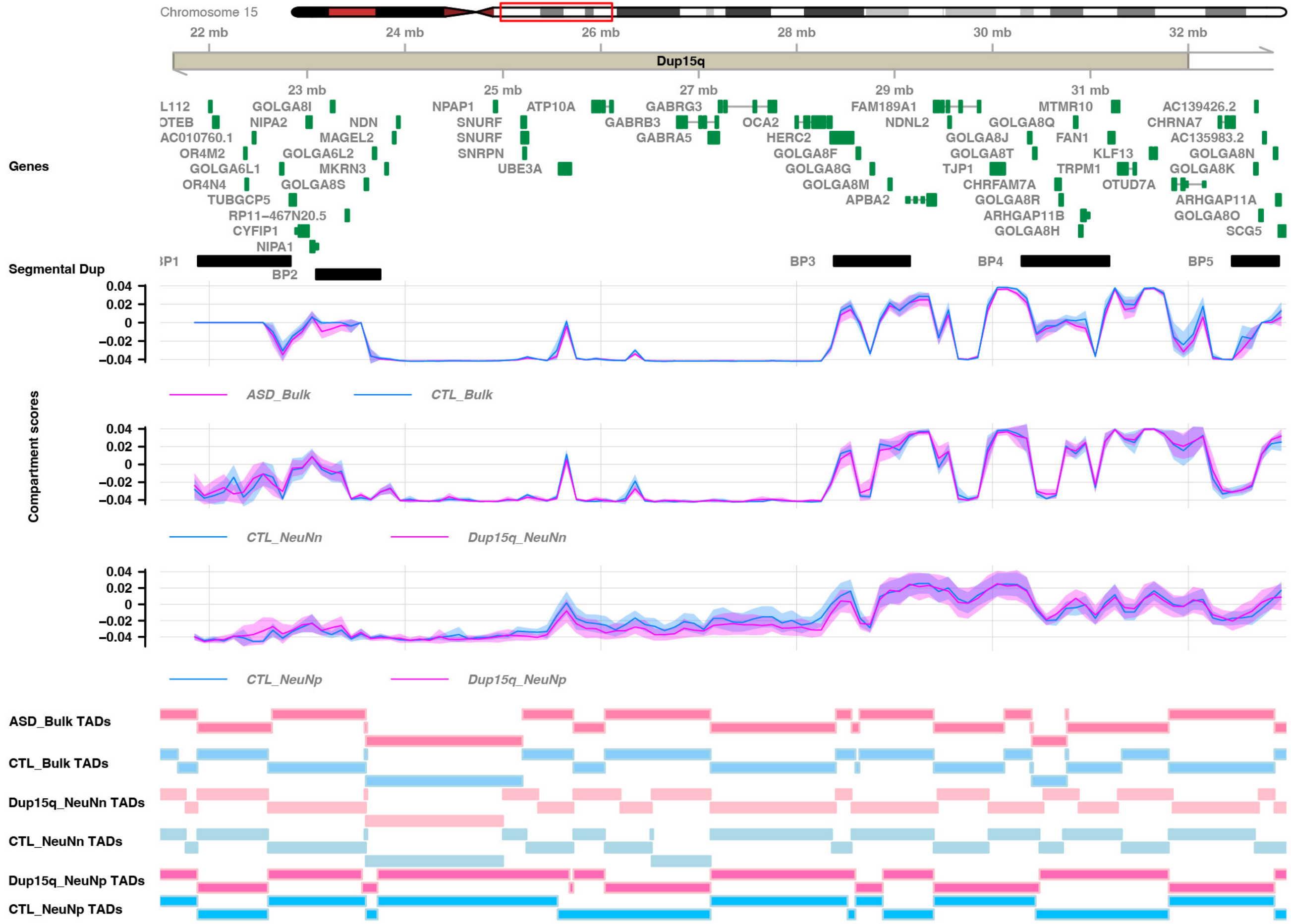

Supplementary Figure 11

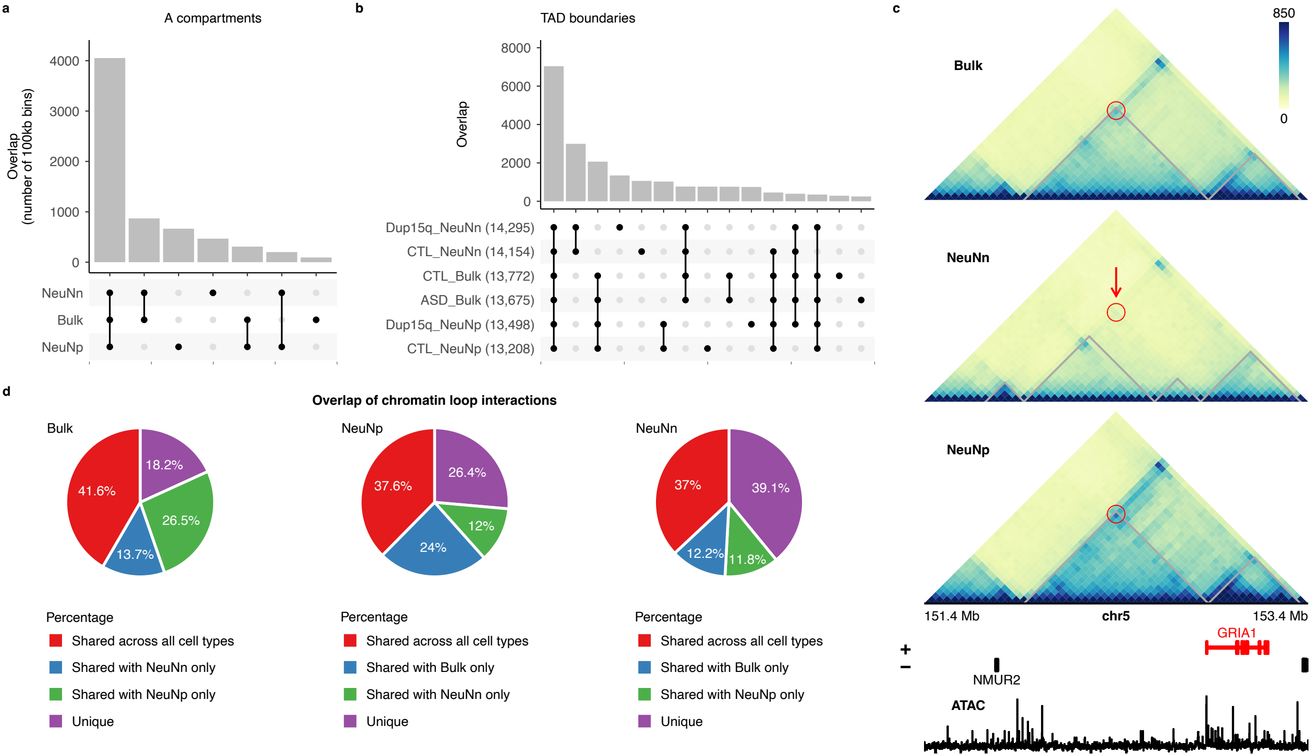

Supplementary Figure 12

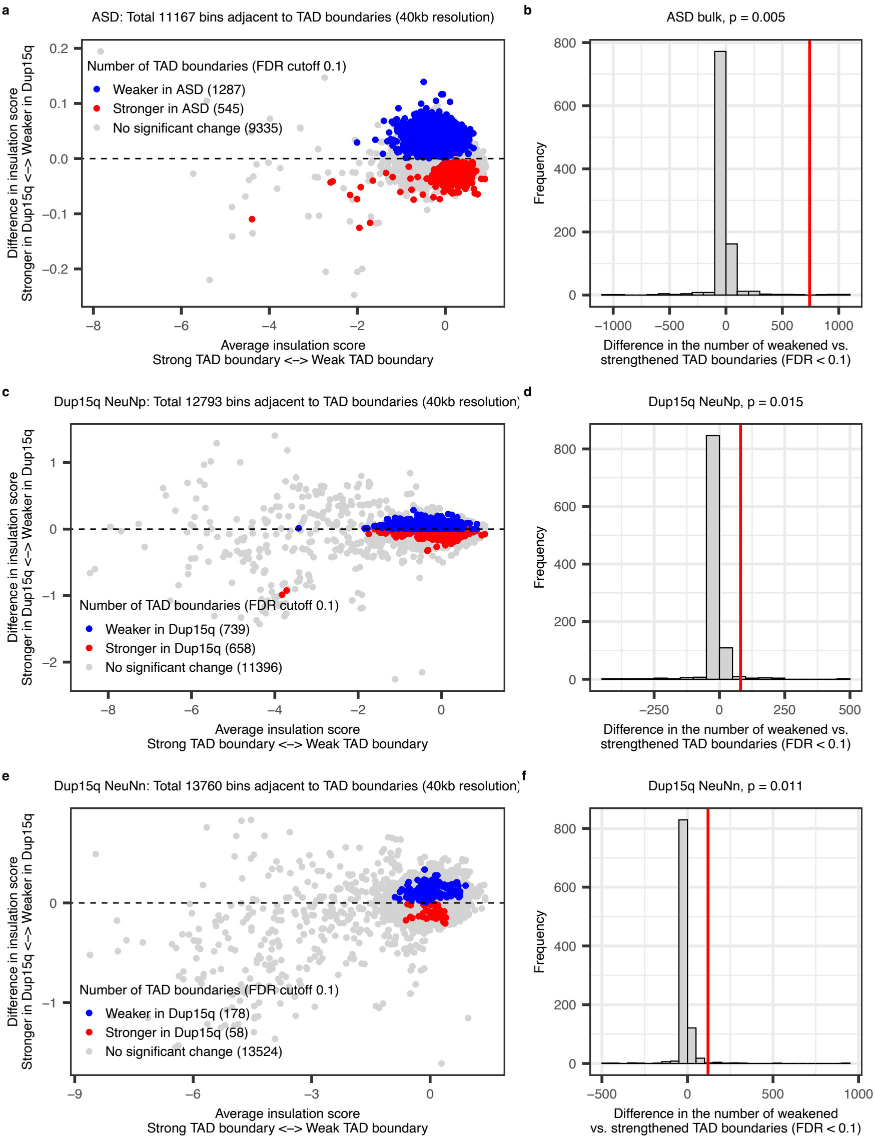

Supplementary Figure 13

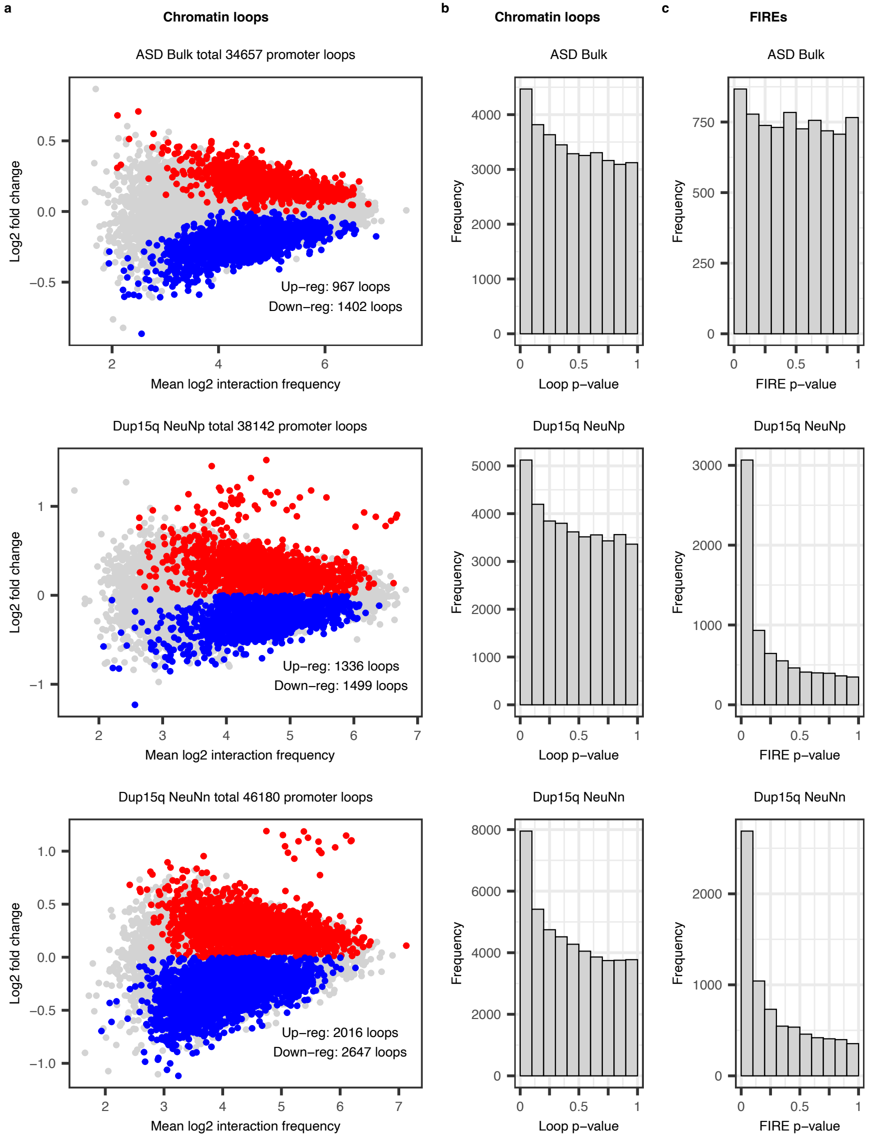

Supplementary Figure 14

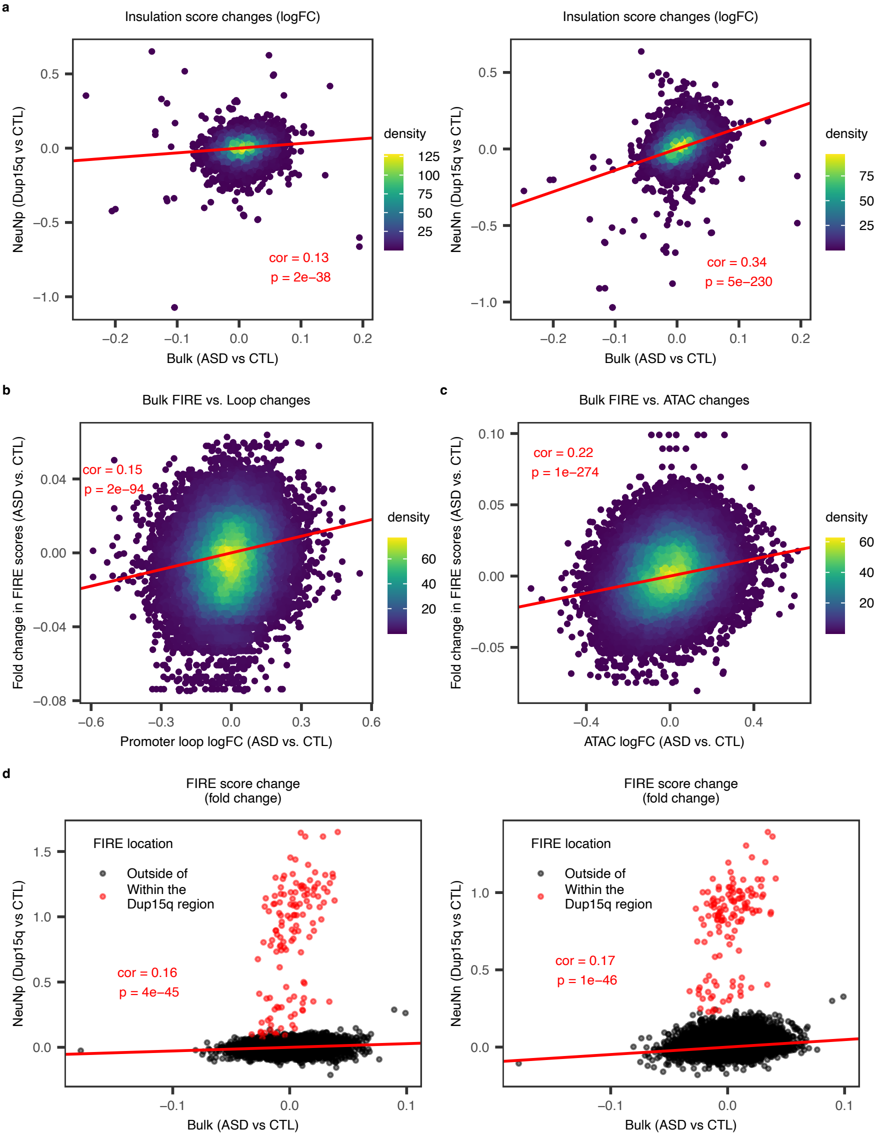

Supplementary Figure 15

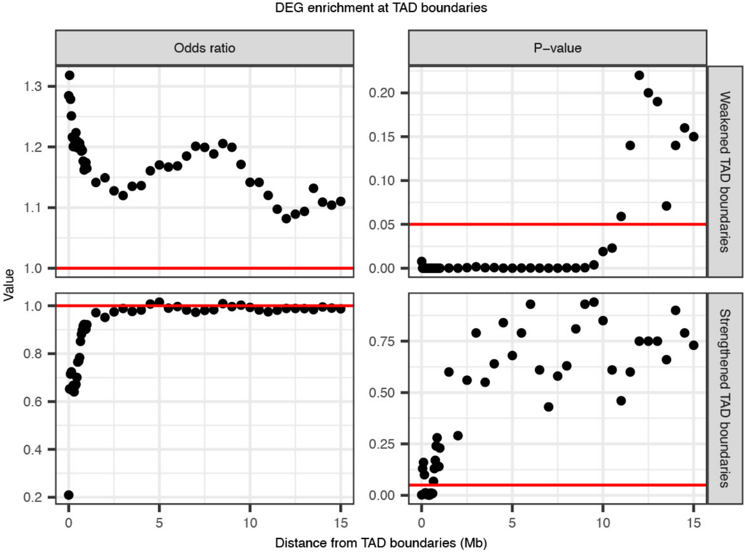
